# Six Month Safety and Efficacy of the BNT162b2 mRNA COVID-19 Vaccine

**DOI:** 10.1101/2021.07.28.21261159

**Authors:** Stephen J. Thomas, Edson D. Moreira, Nicholas Kitchin, Judith Absalon, Alejandra Gurtman, Stephen Lockhart, John L. Perez, Gonzalo Pérez Marc, Fernando P. Polack, Cristiano Zerbini, Ruth Bailey, Kena A. Swanson, Xia Xu, Satrajit Roychoudhury, Kenneth Koury, Salim Bouguermouh, Warren V. Kalina, David Cooper, Robert W. Frenck, Laura L. Hammitt, Özlem Türeci, Haylene Nell, Axel Schaefer, Serhat Ünal, Qi Yang, Paul Liberator, Dina B. Tresnan, Susan Mather, Philip R. Dormitzer, Uğur Şahin, William C. Gruber, Kathrin U. Jansen, C4591001 Clinical Trial Group

**Affiliations:** State University of New York, Upstate Medical University, Syracuse, NY; Associação Obras Sociais Irmã Dulce and Oswaldo Cruz Foundation, Bahia, Brazil; Vaccine Research and Development, Pfizer Inc, Hurley, UK; Vaccine Research and Development, Pfizer Inc, Pearl River, NY; Vaccine Research and Development, Pfizer Inc, Collegeville, PA; iTrials-Hospital Militar Central, Buenos Aires, Argentina; Fundacion INFANT, Buenos Aires, Argentina; Centro Paulista de Investigação Clinica, São Paulo, Brazil; Global Product Development, Pfizer Inc, Peapack, NJ; Cincinnati Children’s Hospital, Cincinnati, OH; Johns Hopkins Bloomberg School of Public Health, Baltimore, MD; BioNTech, Mainz, Germany; Tiervlei Trial Centre, Karl Bremer Hospital, Cape Town, South Africa; Medizentrum Essen Borbeck, Essen, Germany; Hacettepe University, Ankara, Turkey; Worldwide Safety, Safety Surveillance and Risk Management, Pfizer, Inc, Groton, CT; Worldwide Safety, Safety Surveillance and Risk Management, Pfizer, Inc, Collegeville, PA

**Author notes:** Corresponding Author: Judith Absalon, M.D., M.P.H., F.I.D.S.A., Pfizer Inc, 401 North Middletown Road, Pearl River, NY 10965.

## Abstract

**Background:** BNT162b2 is a lipid nanoparticle-formulated, nucleoside-modified RNA vaccine encoding a prefusion-stabilized, membrane-anchored SARS-CoV-2 full-length spike protein. BNT162b2 is highly efficacious against COVID-19 and is currently authorized for emergency use or conditional approval worldwide. At the time of authorization, data beyond 2 months post-vaccination were unavailable.

**Methods:** In an ongoing, placebo-controlled, observer-blinded, multinational, pivotal efficacy study, 44,165 ≥16-year-old participants and 2,264 12-15-year-old participants were randomized to receive 2 doses, 21 days apart, of 30 µg BNT162b2 or placebo. Study endpoints reported here are vaccine efficacy (VE) against laboratory-confirmed COVID-19 and safety data, both up to 6 months post-vaccination.

**Results:** BNT162b2 continued to be safe and well tolerated. Few participants had adverse events leading to study withdrawal. VE against COVID-19 was 91% (95% CI 89.0-93.2) through up to 6 months of follow-up, among evaluable participants and irrespective of previous SARS-CoV-2 infection. VE of 86%-100% was seen across countries and in populations with diverse characteristics of age, sex, race/ethnicity, and COVID-19 risk factors in participants without evidence of previous SARS-CoV-2 infection. VE against severe disease was 97% (95% CI 80.3−99.9). In South Africa, where the SARS-CoV-2 variant of concern, B.1.351 (beta), was predominant, 100% (95% CI 53.5, 100.0) VE was observed.

**Conclusion:** With up to 6 months of follow-up and despite a gradually declining trend in vaccine efficacy, BNT162b2 had a favorable safety profile and was highly efficacious in preventing COVID-19. (ClinicalTrials.gov number, NCT04368728)

## INTRODUCTION

The COVID-19 pandemic continues, with recent estimates of >187 million cases diagnosed and >4 million deaths.^1^ Vaccines are currently available via emergency use authorization or conditional marketing approval pathways.^2-5^ BNT162b2 is a lipid nanoparticle-formulated,^6^ nucleoside-modified RNA^7^ encoding the SARS-CoV-2 full-length spike glycoprotein in a prefusion stabilized conformation.^8^ To date, >350 million BNT162b2 doses have been administered.

We previously reported safety and efficacy data obtained through a median 2 months of post-immunization follow-up from a global phase 1/2/3 trial of BNT162b2 in ≥16-year-olds. Vaccine efficacy (VE) against COVID-19 was 95%. BNT162b2 had a favorable safety profile in diverse populations.^9^ These data formed the basis for BNT162b2 emergency/conditional authorizations globally.^10^ Safety, efficacy, and immunogenicity data from 12-15-year-old participants in this trial have been reported.^11^

Here, we report safety and efficacy findings from a prespecified analysis of the phase 2/3 portion of the trial up to ∼6 months of follow-up.

## METHODS

### Objectives, Participants, Oversight

This randomized, placebo-controlled, observer-blind phase 1/2/3 study assessed BNT162b2 safety, tolerability, efficacy, and immunogenicity in adolescents and adults (NCT04368728). The current report focuses on phase 2/3 safety assessments in ≥16-year-old and prespecified VE assessments in ≥12-year-old participants through up to 6 months post-immunization. Because enrolment of 12-15-year-olds began on October 15, 2020, 6-month post-immunization data are currently unavailable for this age cohort. Shorter duration safety, immunogenicity, and efficacy data for 12-15-year-olds are reported separately;^11^ however, data for this cohort are included in overall VE analyses reported here.

Participants who were healthy or had stable chronic medical conditions were eligible. An active immunocompromising condition or recent immunosuppressive therapy were exclusion criteria. Participants with a COVID-19 medical history were excluded, though evidence of current or prior SARS-CoV-2 infection on laboratory testing of study-obtained samples was not an exclusion. Study responsibilities and ethical conduct are summarized in the **Supplementary Appendix**. Institutional Review Board or Ethics Committee approval was obtained for each site prior to enrollment of any study participant.

### Procedures

Participants were randomized 1:1 by an interactive web-based system to receive 2 intramuscular injections 21 days apart of 30 μg BNT162b2 (0.3-mL volume/dose) or saline placebo. Starting in December 2020, following availability of BNT162b2 under emergency/conditional use authorizations, ≥16-year-old participants who became eligible for COVID-19 vaccination by national/local recommendations were given the option of unblinding. Those who had been randomized to placebo were offered BNT162b2. After unblinding, participants were followed in an open-label study period.

### Safety

Safety endpoints included solicited, prespecified local reactions, systemic events, and antipyretic/pain medication use ≤7 days after receipt of each vaccine or placebo dose (electronic diary reported) and unsolicited adverse events from dose 1 through 1 month after each dose and serious adverse events from dose 1 through 1 and 6 months post-dose 2. Safety data are presented for blinded follow-up and open-label periods.

### Efficacy

BNT162b2 efficacy against laboratory-confirmed COVID-19 with onset ≥7 days post-dose 2 was assessed descriptively in participants without serological or virological evidence of SARS-CoV-2 infection ≤7 days post-dose 2, and in participants with and without evidence of prior infection. Efficacy against severe COVID-19 was also assessed. Lineages of SARS-CoV-2 detected in midturbinate specimens are reported here for COVID-19 cases occurring ≥7 days post-dose 2 in South African participants without evidence of prior infection. Methods for determining SARS-CoV-2 lineages and case definitions for confirmed and severe COVID-19 are summarized. See **Supplementary Appendix**.

### Statistics

Study populations are summarized in **Table S1**. Safety analyses are presented for ≥16-year-olds without known HIV infection who provided informed consent and received ≥1 BNT162b2 or placebo dose. Safety analyses, which are descriptive and not based on formal hypothesis testing, are presented as counts, percentages and associated Clopper-Pearson 95% CIs for adverse events (according to MedDRA terms) and reactogenicity events for each vaccine group. Safety data reported up to March 13, 2021 are summarized here. CIs in this report are not adjusted for multiplicity.

VE analyses during the blinded period are presented for all randomized ≥12-year-olds without known HIV infection who received ≥1 BNT162b2 or placebo dose. VE was estimated by 100×(1-IRR). IRR is the ratio of the rate of confirmed COVID-19 illness per 1000 person-years follow-up in the BNT162b2 group to that in the placebo group. Descriptive analyses of VE and associated 95% CIs used the Clopper and Pearson method, adjusted for surveillance time, which accounts for potential differential follow-up between the 2 groups. Because the percentage of participants who reported symptoms but were missing a valid polymerase-chain-reaction test result was small and slightly higher in the placebo arm, data for these participants were not imputed in the analysis.

The previously reported primary efficacy objective was achieved based on analysis of 170 accrued, evaluable COVID-19 cases (data cut-off: November 14, 2020).^9^ The current report provides updated efficacy analyses conducted on cases accrued up to March 13, 2021.

## RESULTS

### Participants

Between July 27, 2020 and October 29, 2020, 45,441 ≥16-year-olds were screened, and 44,165 randomized at 152 sites (US [n=130], Argentina [n=1], Brazil [n=2], South Africa [n=4], Germany [n=6], Turkey [n=9]) in the phase 2/3 portion of the study. Of these participants, 44,060 were vaccinated with ≥1 dose (BNT162b2, n=22,030; placebo, n=22,030), and 98% received dose 2 (**Fig.1**). During the blinded period, 51% of participants in each group had 4 to <6 months of follow-up post dose 2; 8% of BNT162b2 recipients and 6% of placebo recipients had ≥6 months follow-up post-dose 2. During blinded and open-label periods combined, 55% of BNT162b2 recipients had ≥6 months follow-up post-dose 2. Participants were 49% female, 82% White, 10% Black/African American, and 26% Hispanic/Latinx; median age was 51 years. Thirty-four percent had BMI ≥30.0 kg/m^2^, 21% had ≥1 underlying comorbidity, and 3% had baseline evidence of prior/current SARS-CoV-2 infection (**Tables 1, S2**).

**Figure 1.**
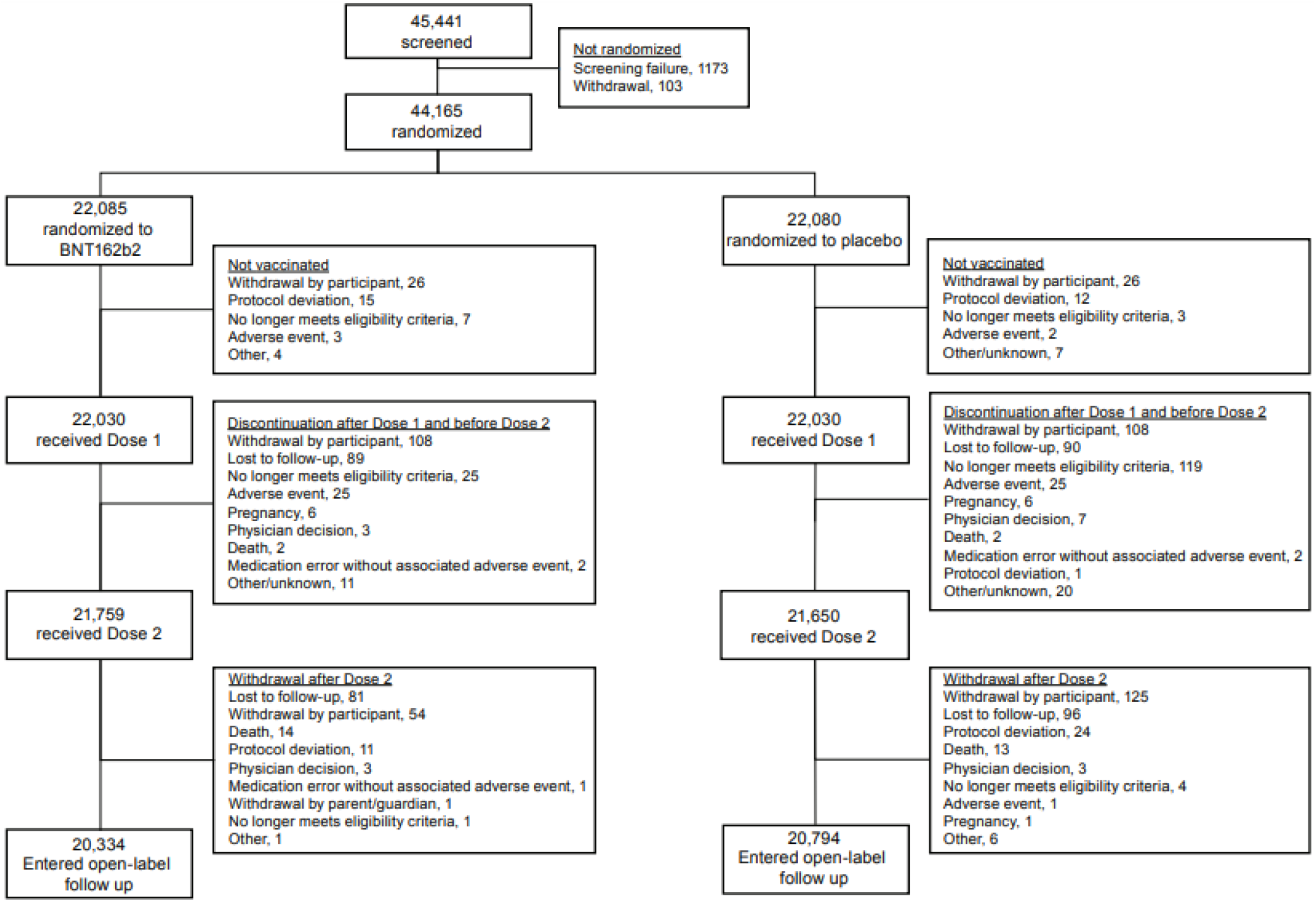
Disposition of Participants. Disposition is presented for all enrolled participants ≥16 years old through the data cut-off (March 13, 2021). Includes 2 deaths in HIV-infected participants (not included in text). Disposition of participants 12-15 years of age has been reported previously.^11^

**Table 1.**
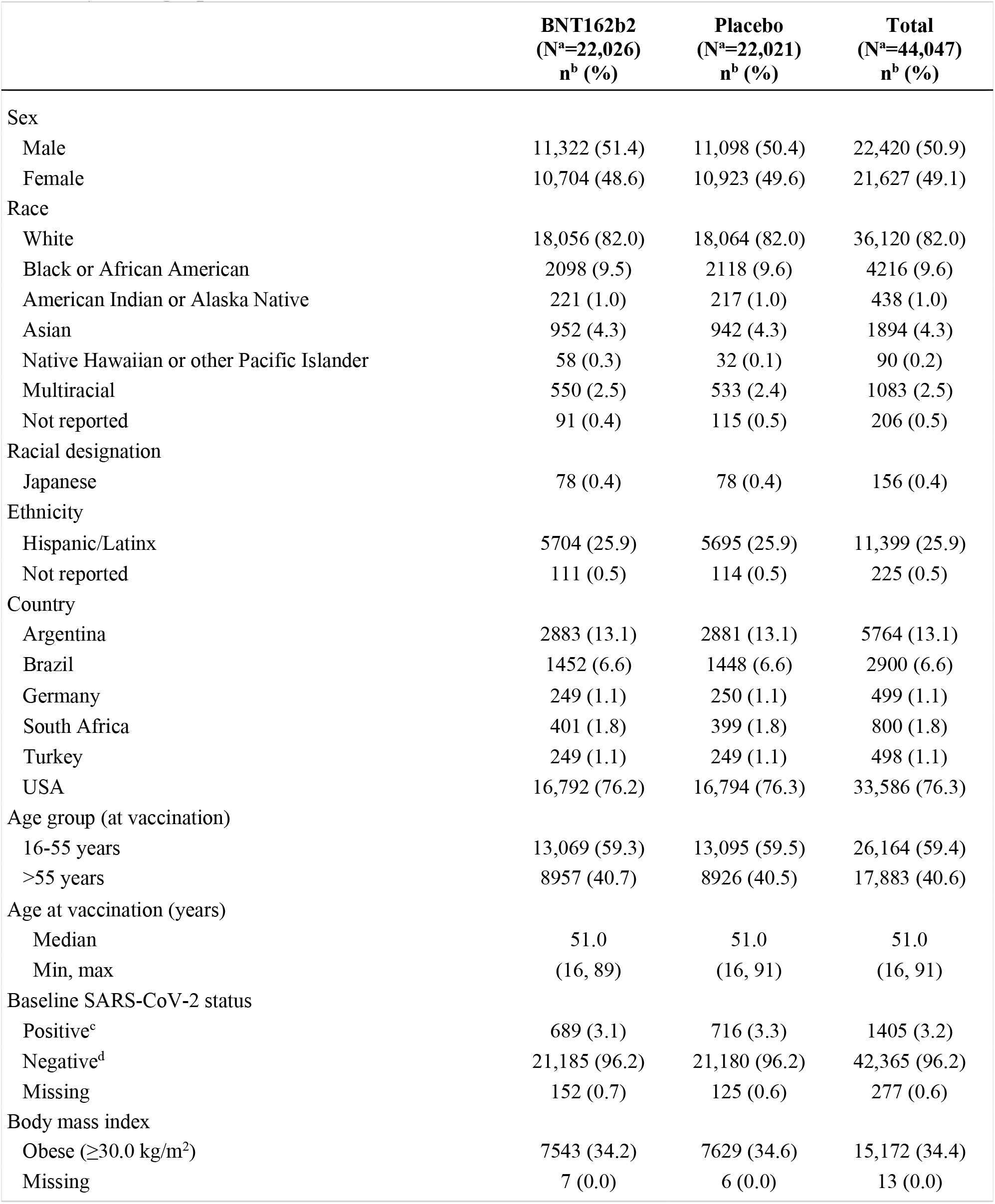
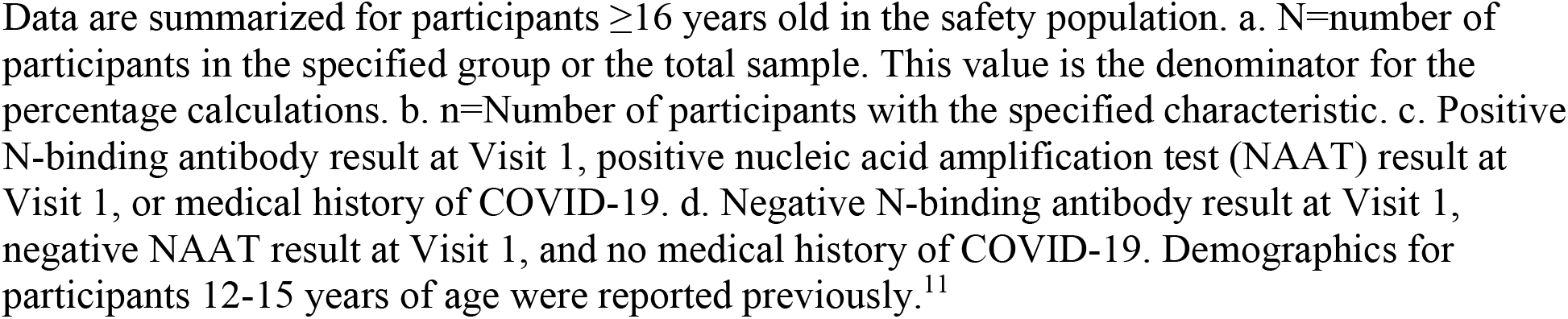
Demographics.

|  | **BNT162b2**<br>(N<sup>a</sup>=22,026)<br>n<sup>b</sup> (%) | **Placebo**<br>(N<sup>a</sup>=22,021)<br>n<sup>b</sup> (%) | **Total**<br>(N<sup>a</sup>=44,047)<br>n<sup>b</sup> (%) |
| --- | --- | --- | --- |
| Sex |  |  |  |
| Male | 11,322 (51.4) | 11,098 (50.4) | 22,420 (50.9) |
| Female | 10,704 (48.6) | 10,923 (49.6) | 21,627 (49.1) |
| Race |  |  |  |
| White | 18,056 (82.0) | 18,064 (82.0) | 36,120 (82.0) |
| Black or African American | 2098 (9.5) | 2118 (9.6) | 4216 (9.6) |
| American Indian or Alaska Native | 221 (1.0) | 217 (1.0) | 438 (1.0) |
| Asian | 952 (4.3) | 942 (4.3) | 1894 (4.3) |
| Native Hawaiian or other Pacific Islander | 58 (0.3) | 32 (0.1) | 90 (0.2) |
| Multiracial | 550 (2.5) | 533 (2.4) | 1083 (2.5) |
| Not reported | 91 (0.4) | 115 (0.5) | 206 (0.5) |
| Racial designation |  |  |  |
| Japanese | 78 (0.4) | 78 (0.4) | 156 (0.4) |
| Ethnicity |  |  |  |
| Hispanic/Latinx | 5704 (25.9) | 5695 (25.9) | 11,399 (25.9) |
| Not reported | 111 (0.5) | 114 (0.5) | 225 (0.5) |
| Country |  |  |  |
| Argentina | 2883 (13.1) | 2881 (13.1) | 5764 (13.1) |
| Brazil | 1452 (6.6) | 1448 (6.6) | 2900 (6.6) |
| Germany | 249 (1.1) | 250 (1.1) | 499 (1.1) |
| South Africa | 401 (1.8) | 399 (1.8) | 800 (1.8) |
| Turkey | 249 (1.1) | 249 (1.1) | 498 (1.1) |
| USA | 16,792 (76.2) | 16,794 (76.3) | 33,586 (76.3) |
| Age group (at vaccination) |  |  |  |
| 16-55 years | 13,069 (59.3) | 13,095 (59.5) | 26,164 (59.4) |
| >55 years | 8957 (40.7) | 8926 (40.5) | 17,883 (40.6) |
| Age at vaccination (years) |  |  |  |
| Median | 51.0 | 51.0 | 51.0 |
| Min, max | (16, 89) | (16, 91) | (16, 91) |
| Baseline SARS-CoV-2 status |  |  |  |
| Positive<sup>c</sup> | 689 (3.1) | 716 (3.3) | 1405 (3.2) |
| Negative<sup>d</sup> | 21,185 (96.2) | 21,180 (96.2) | 42,365 (96.2) |
| Missing | 152 (0.7) | 125 (0.6) | 277 (0.6) |
| Body mass index |  |  |  |
| Obese (≥30.0 kg/m<sup>2</sup>) | 7543 (34.2) | 7629 (34.6) | 15,172 (34.4) |
| Missing | 7 (0.0) | 6 (0.0) | 13 (0.0) |

Data are summarized for participants $\geq 16$ years old in the safety population. a. N=number of participants in the specified group or the total sample. This value is the denominator for the percentage calculations. b. n=Number of participants with the specified characteristic. c. Positive N-binding antibody result at Visit 1, positive nucleic acid amplification test (NAAT) result at Visit 1, or medical history of COVID-19. d. Negative N-binding antibody result at Visit 1, negative NAAT result at Visit 1, and no medical history of COVID-19. Demographics for participants 12-15 years of age were reported previously.<sup>11</sup>

Between October 15, 2020 and January 12, 2021, 2306 12-15-year-olds were screened, 2264 were randomized across 29 US sites, 2260 were vaccinated with ≥1 dose (BNT162b2, n=1131; placebo, n=1129), and 99% received dose 2.^11^ Of vaccinated participants, 58% had ≥2 months follow-up post-dose 2, 49% were female, 86% were White, 4.6% were Black/African American, and 12% were Hispanic/Latinx. Full demographic characteristics are reported elsewhere.^11^

### Safety

#### Reactogenicity

The reactogenicity subset of participants reported here (those using electronic diary for reporting events) includes 9839 ≥16-year-olds. Of this subset, 8183 were included in the previous analysis, and 1656 were enrolled after the data cut-off for that analysis.^9^ The reactogenicity profile for this expanded subset did not differ substantially from that described previously.^9^ The reactogenicity subset included 364 participants with evidence of prior SARS-CoV-2 infection, 9426 without, and 49 who lacked data required to determine prior infection status.

More BNT162b2 than placebo recipients reported local reactions, most commonly mild-to-moderate injection site pain (**Fig. S1A**). Participants with or without evidence of prior SARS-CoV-2 infection reported local reactions with similar frequency and of similar severity. No Grade 4 local reactions were reported.

More BNT162b2 than placebo recipients reported systemic events, most commonly fatigue (**Fig. S1B**). Systemic events were mostly mild-to-moderate, but occasionally severe. Systemic reactogenicity was similar in those with and without evidence of prior SARS-CoV-2 infection, although BNT162b2 recipients with evidence of prior infection reported systemic events more often post-dose 1, and those without evidence reported systemic events more often post-dose 2. For example, 12% of recipients with evidence of prior SARS-CoV-2 infection and 3% of those without reported fever post-dose 1; 8% of those with evidence of prior infection and 15% of those without reported fever post-dose 2. The highest temperature reported was a transient fever >40.0°C on day 2 post-dose 2 in a BNT162b2 recipient without evidence of prior infection.

#### Adverse events

Adverse event analyses during the blinded period are provided for 43,847 ≥16-year-olds (**Table S3**). Reactogenicity events among participants not in the reactogenicity subset are reported as adverse events, resulting in imbalances in adverse events (30% vs 14%), related adverse events (24% vs 6%), and severe adverse events (1.2% vs 0.7%) between BNT162b2 and placebo groups. Decreased appetite, lethargy, asthenia, malaise, night sweats, and hyperhidrosis were new adverse events attributable to BNT162b2 not previously identified in earlier reports. Few participants had serious adverse events or adverse events leading to study withdrawal. No new serious adverse events assessed as related by investigators were reported after data cut-off for the previous report.^9^

Cumulative safety follow-up was available up to 6 months post-dose 2 from combined blinded and open-label periods for 12,006 participants originally randomized to BNT162b2. The longer follow-up for this report, including open-label observation of original BNT162b2 recipients and placebo recipients who received BNT162b2 after unblinding, revealed no new safety signals relative to the previous report.^9^

During the blinded, controlled period, 15 BNT162b2 and 14 placebo recipients died; during the open-label period, 3 BNT162b2 and 2 original placebo recipients who received BNT162b2 after unblinding died. None of these deaths were considered related to BNT162b2 by investigators. Causes of death were balanced between BNT162b2 and placebo groups **(Table S4)**.

Safety monitoring will continue per protocol for 2 years post-dose 2 for participants who originally received BNT162b2 and for 18 months after the second BNT162b2 dose for placebo recipients who received BNT162b2 after unblinding.

### Efficacy

Among 42,094 evaluable ≥12-year-olds without evidence of prior SARS-CoV-2 infection, 77 COVID-19 cases with onset ≥7 days post-dose 2 were observed through the data cut-off (March 13, 2021) among vaccine recipients and 850 among placebo recipients, corresponding to 91.3% VE (95% CI [89.0-93.2]; **Table 2**). Among 44,486 evaluable participants, irrespective of prior SARS-CoV-2 infection, 81 COVID-19 cases were observed among vaccine and 873 among placebo recipients, corresponding to 91.1% VE (95% CI [88.8-93.0]).

**Table 2.** Vaccine Efficacy against COVID-19 from 7 Days after Dose 2 During the Blinded Placebo-Controlled Follow-up Period (Evaluable Efficacy Population, ≥12 Years Old)

| Efficacy Endpoint | BNT162b2 |  | Placebo |  | VE (%) | (95% CI<sup>d</sup>) | Posterior Probability (VE >30% \| data)<sup>e</sup> |
| --- | --- | --- | --- | --- | --- | --- | --- |
|  | n1<sup>a</sup> | Surveillance Time<sup>b</sup> (n2<sup>c</sup>) | n1<sup>a</sup> | Surveillance Time<sup>b</sup> (n2<sup>c</sup>) |  |  |  |
| COVID-19 occurrence from 7 days after dose 2 in participants without prior evidence of infection | 77 | (N<sup>f</sup>=20,998) | 850 | (N<sup>f</sup>=21,096) | 91.3 | (89.0, 93.2) | >0.9999 |
|  |  | 6.247 (20,712) |  | 6.003 (20,713) |  |  |  |
| COVID-19 occurrence from 7 days after dose 2 in participants with and those without prior evidence of infection | 81 | (N<sup>f</sup>=22,166) | 873 | (N<sup>f</sup>=22,320) | 91.1 | (88.8, 93.0) | >0.9999 |
|  |  | 6.509 (21,642) |  | 6.274 (21,689) |  |  |  |

VE=vaccine efficacy. Participants who had no serological or virological evidence (prior to 7 days after receipt of the last dose) of past SARS-CoV-2 infection (ie, N-binding antibody [serum] negative at Visit 1 and SARS-CoV-2 not detected by NAAT [nasal swab] at Visits 1 and 2), and had negative NAAT (nasal swab) at any unscheduled visit prior to 7 days post-dose 2 were included in the analysis. a. n1=number of participants meeting the endpoint definition. b. Total surveillance time in 1000 person-years for the given endpoint across all participants within each group at risk for the endpoint. Time period for COVID-19 case accrual is from 7 days after dose 2 to the end of the surveillance period. c. n2=number of participants at risk for the endpoint. d. CI for VE was derived based on the Clopper and Pearson method adjusted for surveillance time. e. Posterior probability was calculated using a beta-binomial model with prior beta (0.700102, 1) adjusted for surveillance time. f. N=number of participants in the specified group; total population without prior baseline infection N=42,094; total population with and without prior infection N=44,486.

In the all-available population with evidence of prior SARS-CoV-2 infection based on positive baseline N-binding antibody test, 2 COVID-19 cases were observed post-dose 1 among vaccine and 7 among placebo recipients. In participants with evidence of SARS-CoV-2 infection by positive nucleic acid amplification test at baseline, no difference in COVID-19 cases was observed between vaccine (n=10) and placebo (n=9) recipients (**Table S5**). COVID-19 was less frequent among placebo recipients with positive N-binding antibodies at study entry (7/542; ∼1.3% attack rate) than among those without evidence of infection at study entry (1015/21,521; ∼4.7% attack rate), indicating ∼72.6% protection by previous infection.

From dose 1 to before dose 2, 46 COVID-19 cases were observed in BNT162b2 and 110 in placebo recipients (with or without evidence of prior infection), corresponding to 58.4% VE (95% CI [40.8-71.2]) (**Fig. 2**). During the interval from the approximate start of observed protection at 11 days post-dose 1 to before dose 2, VE increased to 91.7% (95% CI 79.6-97.4). From its peak post-dose 2, observed VE declined. From 7 days to <2 months post-dose 2, VE was 96.2% (95% CI [93.3-98.1]); from 2 months to <4 months, VE was 90.1% (95% CI [86.6-92.9]); and from 4 months to the data cut-off, VE was 83.7% (95% CI [74.7-89.9]).

**Figure 2.**
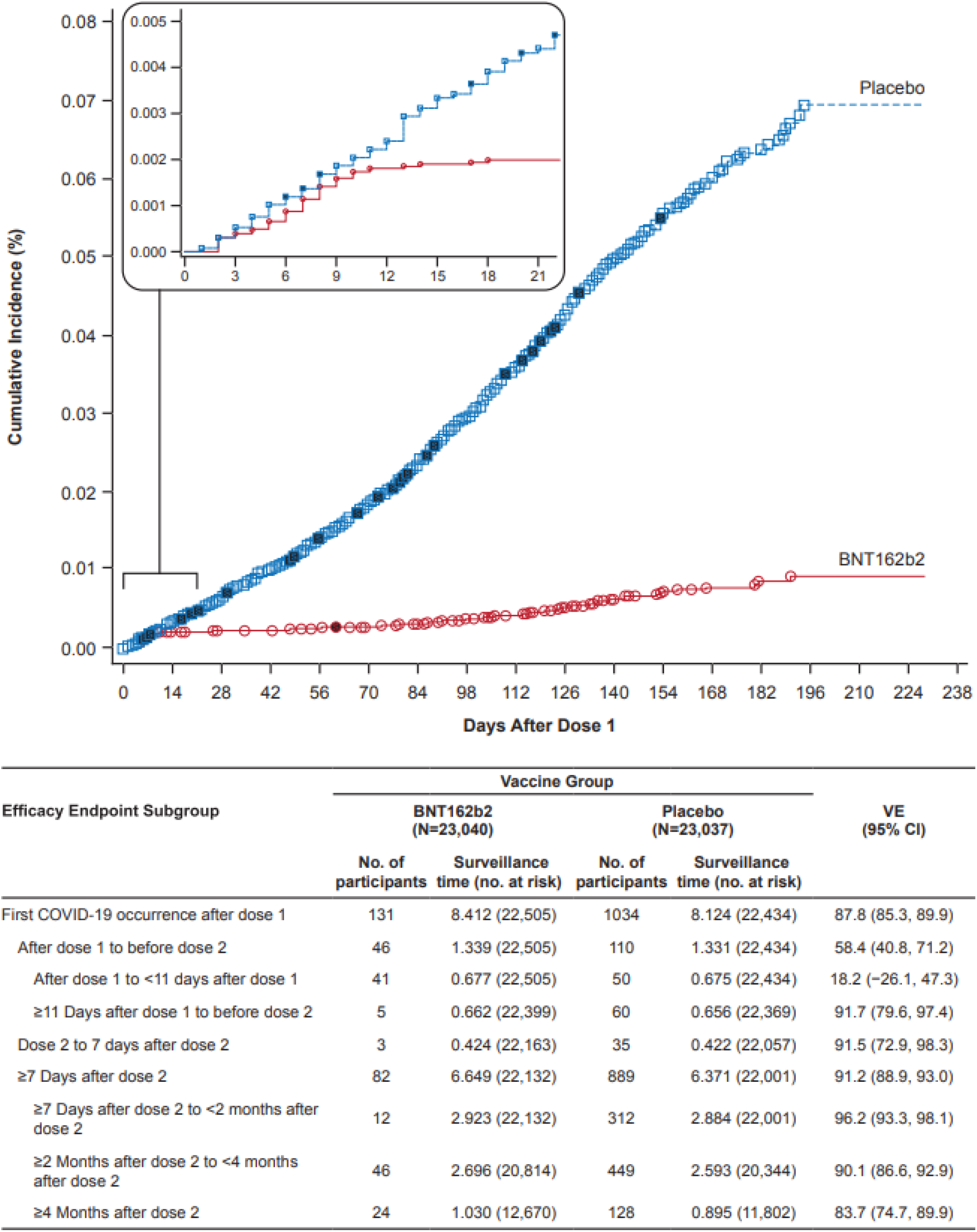
Efficacy of BNT162b2 against COVID-19 Occurrence after Dose 1 During the Blinded Placebo-controlled Follow-up Period. Cumulative incidence curve of first COVID-19 occurrence after dose 1 (all-available efficacy population; ≥12 years of age). VE=vaccine efficacy. Each symbol represents COVID-19 cases starting on a given day, and filled symbols represent severe COVID-19 cases. Due to overlapping dates, some symbols represent more than one case. The inset shows the same data on an enlarged y axis, through 21 days. Total surveillance time in 1000 person-years is for the given endpoint across all participants within each group at risk for the endpoint. Time period for COVID-19 case accrual is from dose 1 to the end of the surveillance period for the overall row and from the start to the end of the range stated for each time interval. CI for VE was derived based on the Clopper and Pearson method adjusted for surveillance time.

Of 31 cases of severe, FDA-defined COVID-19,^12^ with onset post-dose 1, 30 occurred in placebo recipients, corresponding to 96.7% VE (95% CI 80.3-99.9) against severe COVID-19 (**Fig. 2, Table S6**).

Although the study was not powered to definitively assess efficacy by subgroup, supplemental analyses indicated that VE post-dose 2 among subgroups defined by age, sex, race, ethnicity, presence of comorbid conditions, and country was generally consistent with that observed in the overall population (**Tables 3, S7**).

**Table 3.**
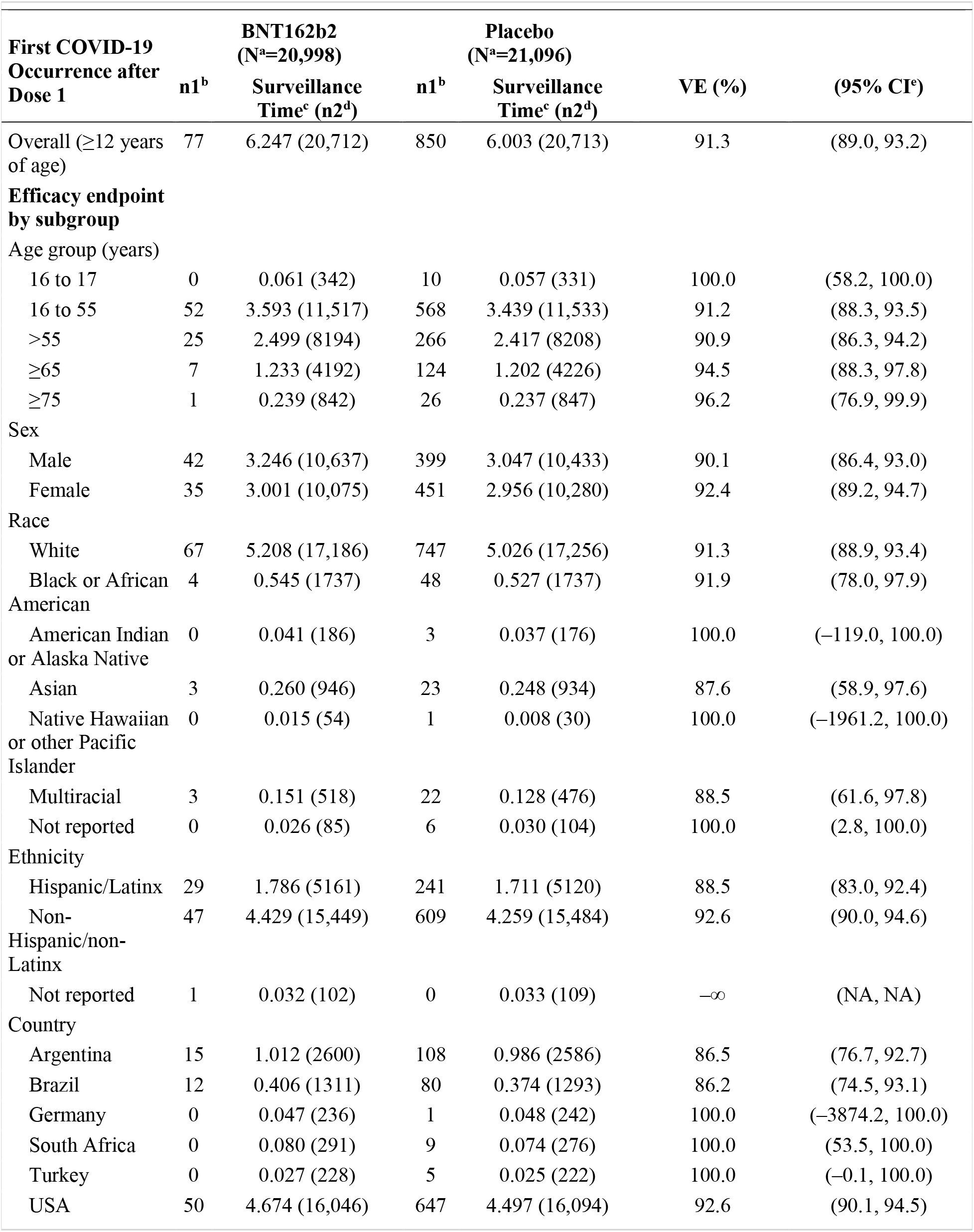
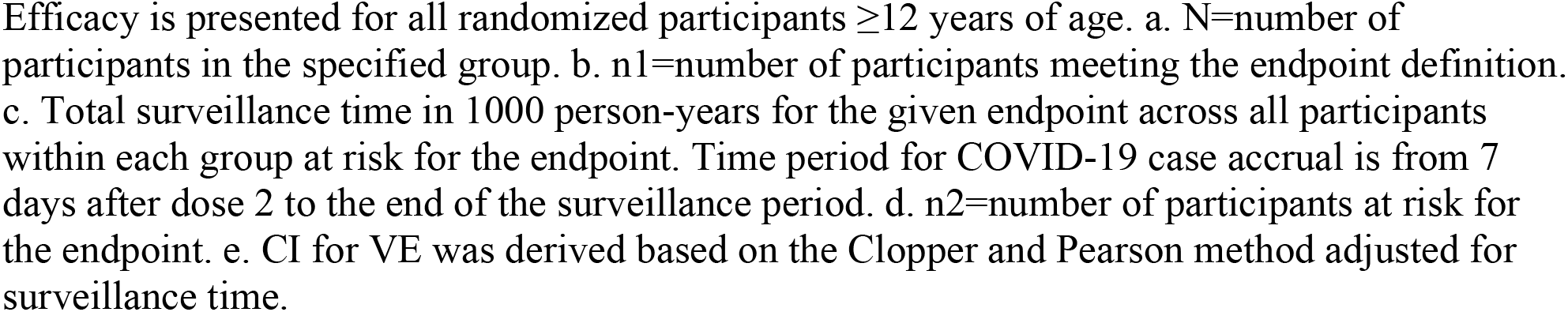
Vaccine Efficacy Overall and by Subgroup in Participants Without Evidence of Infection Prior to 7 Days After Dose 2 During the Blinded Placebo Controlled Follow-up Period.

| First COVID-19 Occurrence after Dose 1 | BNT162b2 (N<sup>a</sup>=20,998) |  | Placebo (N<sup>a</sup>=21,096) |  | VE (%) | (95% CI<sup>e</sup>) |
| --- | --- | --- | --- | --- | --- | --- |
|  | n1<sup>b</sup> | Surveillance Time<sup>c</sup> (n2<sup>d</sup>) | n1<sup>b</sup> | Surveillance Time<sup>c</sup> (n2<sup>d</sup>) |  |  |
| Overall (≥12 years of age) | 77 | 6.247 (20,712) | 850 | 6.003 (20,713) | 91.3 | (89.0, 93.2) |
| **Efficacy endpoint by subgroup** |  |  |  |  |  |  |
| Age group (years) |  |  |  |  |  |  |
| 16 to 17 | 0 | 0.061 (342) | 10 | 0.057 (331) | 100.0 | (58.2, 100.0) |
| 16 to 55 | 52 | 3.593 (11,517) | 568 | 3.439 (11,533) | 91.2 | (88.3, 93.5) |
| >55 | 25 | 2.499 (8194) | 266 | 2.417 (8208) | 90.9 | (86.3, 94.2) |
| ≥65 | 7 | 1.233 (4192) | 124 | 1.202 (4226) | 94.5 | (88.3, 97.8) |
| ≥75 | 1 | 0.239 (842) | 26 | 0.237 (847) | 96.2 | (76.9, 99.9) |
| Sex |  |  |  |  |  |  |
| Male | 42 | 3.246 (10,637) | 399 | 3.047 (10,433) | 90.1 | (86.4, 93.0) |
| Female | 35 | 3.001 (10,075) | 451 | 2.956 (10,280) | 92.4 | (89.2, 94.7) |
| Race |  |  |  |  |  |  |
| White | 67 | 5.208 (17,186) | 747 | 5.026 (17,256) | 91.3 | (88.9, 93.4) |
| Black or African American | 4 | 0.545 (1737) | 48 | 0.527 (1737) | 91.9 | (78.0, 97.9) |
| American Indian or Alaska Native | 0 | 0.041 (186) | 3 | 0.037 (176) | 100.0 | (-119.0, 100.0) |
| Asian | 3 | 0.260 (946) | 23 | 0.248 (934) | 87.6 | (58.9, 97.6) |
| Native Hawaiian or other Pacific Islander | 0 | 0.015 (54) | 1 | 0.008 (30) | 100.0 | (-1961.2, 100.0) |
| Multiracial | 3 | 0.151 (518) | 22 | 0.128 (476) | 88.5 | (61.6, 97.8) |
| Not reported | 0 | 0.026 (85) | 6 | 0.030 (104) | 100.0 | (2.8, 100.0) |
| Ethnicity |  |  |  |  |  |  |
| Hispanic/Latinx | 29 | 1.786 (5161) | 241 | 1.711 (5120) | 88.5 | (83.0, 92.4) |
| Non-Hispanic/non-Latinx | 47 | 4.429 (15,449) | 609 | 4.259 (15,484) | 92.6 | (90.0, 94.6) |
| Not reported | 1 | 0.032 (102) | 0 | 0.033 (109) | —∞ | (NA, NA) |
| Country |  |  |  |  |  |  |
| Argentina | 15 | 1.012 (2600) | 108 | 0.986 (2586) | 86.5 | (76.7, 92.7) |
| Brazil | 12 | 0.406 (1311) | 80 | 0.374 (1293) | 86.2 | (74.5, 93.1) |
| Germany | 0 | 0.047 (236) | 1 | 0.048 (242) | 100.0 | (-3874.2, 100.0) |
| South Africa | 0 | 0.080 (291) | 9 | 0.074 (276) | 100.0 | (53.5, 100.0) |
| Turkey | 0 | 0.027 (228) | 5 | 0.025 (222) | 100.0 | (-0.1, 100.0) |
| USA | 50 | 4.674 (16,046) | 647 | 4.497 (16,094) | 92.6 | (90.1, 94.5) |

Efficacy is presented for all randomized participants $\geq 12$ years of age. a. N=number of participants in the specified group. b. n1=number of participants meeting the endpoint definition. c. Total surveillance time in 1000 person-years for the given endpoint across all participants within each group at risk for the endpoint. Time period for COVID-19 case accrual is from 7 days after dose 2 to the end of the surveillance period. d. n2=number of participants at risk for the endpoint. e. CI for VE was derived based on the Clopper and Pearson method adjusted for surveillance time.

Given concern about SARS-CoV-2 lineage B.1.351 (beta), which appears less efficiently neutralized by BNT162b2 immune sera than many other lineages,^13^ whole viral genome sequencing was performed on midturbinate samples from COVID-19 cases observed in South Africa, where this lineage is prevalent. Nine COVID-19 cases were observed in South African participants without evidence of prior SARS-CoV-2 infection, all in placebo recipients, corresponding to 100% VE (95% CI 53.5-100.0; **Table 3**). Midturbinate specimens from 8/9 cases contained sufficient viral RNA for whole genome sequencing. All viral genomes were of the B.1.351 lineage (GISAID accession codes in **Supplementary Appendix**).

## DISCUSSION

In this update to the preliminary safety and efficacy report of a 2-dose regimen of 30 μg BNT162b2 given 21 days apart, 91.1% VE against COVID-19 was observed from 7 days to 6 months post-dose 2 in ≥12-year-old participants. VE against severe disease with onset post-dose 1 was ∼97%. This finding, combined with the totality of available evidence, including real-world effectiveness data,^14-17^ alleviates theoretical concerns over potential vaccine-mediated disease enhancement.^18^

The benefit of BNT162b2 immunization started ∼11 days post-dose 1, with 91.7% VE from 11 days post-dose 1 until dose 2. The study cannot provide information on persistence of protection after a single dose, as 98.5% of participants received dose 2 as scheduled during the blinded study period. As recently reported, although non-neutralizing viral antigen-binding antibody levels rise between the first and second BNT162b2 dose, serum neutralizing titers are low or undetectable during this interval.^19^ Early protection against COVID-19 without robust serum neutralization indicates that neutralizing titers alone do not appear to explain early BNT162b2-mediated protection from COVID-19. Other immune mechanisms (e.g., innate immune responses, CD4^+^ or CD8^+^ T-cell responses, B-cell memory responses, antibody-dependent cytotoxicity) may contribute to protection.^20-25^

Efficacy peaked at 96.2% during the interval from 7 days to <2 months post-dose 2, and declined gradually to 83.7% from 4 months post-dose 2 to the data cut-off, an average decline of ∼6% every 2 months. Ongoing follow-up is needed to understand persistence of the vaccine effect over time, the need for booster dosing, and timing of such a dose. Most participants who initially received placebo have now been immunized with BNT162b2, ending the placebo-controlled part of the study. Nevertheless, ongoing observation of participants through up to 2 years in this study, together with real-world effectiveness data,^14-17^ will determine whether a booster is likely to be beneficial after a longer interval. Booster trials to evaluate safety and immunogenicity of BNT162b2 are underway to prepare for this possibility.

From 7 days post-dose 2, 86%-100% efficacy was observed across diverse demographic profiles, including age, sex, race/ethnicity and factors increasing risk of COVID-19, such as high BMI, and other comorbidities. BNT162b2 was also highly efficacious in various geographic regions including North America, Europe, South Africa and Latin America. Although VE was slightly lower in Latin American countries, BNT162b2 had high efficacy of ∼86% in Argentina and Brazil. Circulation of SARS-CoV-2 variants, some of which are associated with more rapid transmission, and potentially, greater pathogenicity,^26^ has raised concerns that such variants could evade vaccine-mediated protection. Our studies of in vitro neutralization of a variety of SARS-CoV-2 variants have, to date, found that all tested BNT162b2-immune sera neutralize all tested variants.^13,27-31^ The variant with the greatest reduction in neutralization, B.1.351, which has been the dominant South African strain, is still neutralized at serum titers higher than those observed at the onset of protection against COVID-19 after the first vaccine dose.^9,13,19^ Consistent with this finding, BNT162b2 had 100% (95% CI 53.5, 100.0) observed efficacy against COVID-19 in South Africa (placebo recipients, 9 cases; BNT162b2 recipients, 0 cases), and 8/9 cases for which sequence information could be obtained were B.1.351 lineage SARS-CoV-2.

Safety data are now available for ∼44,000 ≥16-year-old participants; 12,006 individuals have ≥6 months of safety follow-up data after a second BNT162b2 dose. The safety profile observed at a median of 2 months post-immunization was confirmed up to 6 months post-immunization in this analysis. No cases of myocarditis were noted.

Before immunization, 3% of ≥16-year-olds had evidence of SARS-CoV-2 infection. Although this group experienced slightly higher systemic reactogenicity events post-dose 1 than those without evidence of prior infection, the group experienced slightly less reactogenicity post-dose 2 than those without prior infection. Thus, there was minimal observed difference in the overall reactogenicity profile based on baseline infection status. Nine COVID-19 cases were observed among individuals with previous serologically defined natural infection: 2 cases in vaccine and 7 in placebo recipients. These data support the current practice of immunizing without screening for evidence of prior infection.

This report has several limitations. Duration of protection and safety data that could be collected in a blinded, placebo-controlled manner were limited by the ethical and practical need to immunize eligible initial placebo recipients under EUA and according to public health authority recommendations. Data presented here do not address whether vaccination prevents asymptomatic infection, but evaluation of that question is ongoing in this study, and real-world data suggest that BNT162b2 prevents asymptomatic infection.^32,33^ Preliminary analyses of breakthrough cases have not yet identified a correlate of protection, as vaccine protection rates remain high. This report does not address VE and safety in pregnant women and in children younger than 12 years. Studies evaluating BNT162b2 in these populations are ongoing.

The data in this report demonstrate that BNT162b2 prevents COVID-19 effectively for up to 6 months post-dose 2 across diverse populations, despite the emergence of SARS-CoV-2 variants, including the B.1.351 lineage, and the vaccine continues to show a favorable safety profile.

## Data Availability

Upon request, and subject to review, Pfizer will provide the data that support the findings of this study. Subject to certain criteria, conditions, and exceptions, Pfizer may also provide access to the related individual anonymized participant data. See https://www.pfizer.com/science/clinical-trials/trial-data-and-results for more information.

## Acknowledgements

We would like to thank all the participants who volunteered for this study.

We would also like to acknowledge the contributions of the C4591001 Clinical Trial Group (See supplemental appendix for full list).

The authors would like to thank Jonathan Zenilman, MD (Chair), Robert Belshe, MD, Kathryn Edwards, MD, Stephen Self, PhD and Lawrence Stanberry, MD, members of the C4591001 Data Safety Monitoring Board for their dedication and their diligent review of the data.

The first draft of the manuscript was written by Judith Absalon. The authors would like to thank Tricia Newell, Sheena Hunt, and Philippa Jack of ICON plc (North Wales, PA) for editorial support, which was funded by Pfizer.

Pfizer Inc: Greg Adams, Negar Aliabadi, Mohanish Anand, Fred Angulo, Ayman Ayoub, Melissa Bishop-Murphy, Mark Boaz, Christopher Bowen, Donna Boyce, Sarah Burden, Andrea Cawein, Patrick Caubel, Darren Cowen, Kimberly Ann Cristall, Michael Cruz, Daniel Curcio, Gabriela Dávila, Carmel Devlin, Gokhan Duman, Niesha Foster, Maja Gacic, Juleen Gayed, Ahmed Hassan, Luis Jodar, Stephen Kay, William Lam, Esther Ladipo, Joaquina Maria Lazaro, Marie-Pierre Hellio Le Graverand-Gastineau, Kwok Lee, Zhenghui Li, Jacqueline Lowenberg, Hua Ma, Rod MacKenzie, Robert Maroko, Jason McKinley, Tracey Mellelieu, Neda Aghajani Memar, Farheen Muzaffar, Brendan O’Neill, Jason Painter, Elizabeth Paulukonis, Allison Pfeffer, Katie Puig, Kimberly Rarrick, Balaji Prabu Raja, Christine Rainey, Kellie Lynn Richardson, Elizabeth Rogers, Melinda Rottas, Charulata Sabharwal, Uzma Sarwar, Vilas Satishchandran, Harpreet Seehra, Judy Sewards, Huiqing Si, Helen Smith, David Swerdlow, James Trammel, Elisa Harkins Tull, Sarah Tweedy, Erica Weaver, John Wegner, Jenah West, Christopher Webber, David C Whritenour, Fae Wooding, Emily Worobetz, Nita Zalavadia, Liping Zhang, the Vaccines Clinical Assay Team, the Vaccines Assay Development Team and all of the Pfizer colleagues not named here who contributed to the success of this study.

BioNTech: Corinna Rosenbaum, Christian Miculka, Andreas Kuhn, Ferdia Bates, Paul Strecker, Ruben Rizzi, Martin Bexon, Eleni Lagkadinou, and Alexandra Kemmer-Brück.

Polymun: Dietmar Katinger and Andreas Wagner

