## Supplementary Appendix for "Six Month Safety and Efficacy of the BNT162b2 mRNA COVID-19 Vaccine"

### List of Investigators

| Name | Institute | Location |
|---|---|---|
| Aberg, Judith | Icahn School of Medicine at Mount Sinai | New York, NY, USA |
| Addo, Marylyn | Universitätsklinikum Hamburg-Eppendorf | Hamburg, Germany |
| Akhan, Sıla | Kocaeli University Med | Kocaeli, Turkey |
| Albertson, Timothy | University of California, Davis | Sacramento, CA, USA |
| Al-Ibrahim, Mohamed | SNBL Clinical Pharmacology Center | Baltimore, MD, USA |
| Altın, Sedat | Yedikule Chest Diseases and Thoracic Surgery Training and Research Hospital | Istanbul, Turkey |
| Anderson, Corey | Clinical Research Consortium | Tempe, AZ, USA |
| Andrews, Charles | Diagnostics Research Group | San Antonio, TX, USA |
| Arora, Samir | Aventiv Research | Columbus, OH, USA |
| Balik, Ismail | Ankara University Medical School | Ankara, Turkey |
| Barnett, Elizabeth | Boston Medical Center | Boston, MA, USA |
| Bauer, George | Benchmark Research | Metairie, LA, USA |
| Baumann-Noss, Sybille | CRS Clinical Research Services Berlin | Berlin, Germany |
| Berhe, Mezgebe | North Texas Infectious Diseases Consultants | Dallas, TX, USA |
| Bradley, Paul | Meridian Clinical Research | Savannah, GA, USA |
| Brandon, Donald | California Research Foundation | San Diego, CA, USA |
| Brune, Daniel | Optimal Research | Peoria, IL, USA |
| Burgher, Abram | The Pain Center of Arizona | Peoria, AZ, USA |
| Butcher, Bain | Sterling Research Group | Cincinnati, OH, USA |
| Butuk, David | Solaris Clinical Research | Meridian, ID, USA |
| Cannon, Kevin | PMG Research of Wilmington | Wilmington, NC, USA |
| Cardona, Jose | Indago Research and Health Center | Hialeah, FL, USA |
| Cavelli, Rachael | State University of New York Upstate Medical University | Syracuse, NY, USA |
| Chalhoub, Fadi | CNS Healthcare | Jacksonville, FL, USA |
| Christensen, Shane | J. Lewis Research | Salt Lake City, UT, USA |
| Christensen, Tom | Waterway Family Medicine | Little River, SC, USA |
| Chu, Laurence | Benchmark Research | Austin, TX, USA |
| Cox, Steven | Lynn Institute of Norman | Norman, OK, USA |
| Crook, Gretchen | Austin Regional Clinic: ARC Wilson Parke | Austin, TX, USA |
| Davis, Matthew | Rochester Clinical Research | Rochester, NY, USA |
| Davit, Rajesh | Sterling Research Group | Cincinnati, OH, USA |
| Denham, Douglas | Clinical Trials of Texas (CTT) | San Antonio, TX, USA |
| Dever, Michael | Clinical Neuroscience Solutions | Orlando, FL, USA |
| Donskey, Curtis | Louis Stokes Cleveland VA Medical Center | Cleveland, OH, USA |
| Doust, Matthew | Hope Research Institute | Phoenix, AZ, USA |
| Dunn, Michael | Quality Clinical Research | Omaha, NE, USA |
| Earl, John | PMG Research of Hickory | Hickory, NC, USA |
| Eder, Frank | United Medical Associates - Meridian Clinical Research | Binghamton, NY, USA |
| Eich, Andreas | IKF Pneumologie | Frankfurt, Germany |
| Ensz, David | Meridian Clinical Research | Dakota Dunes, SD, USA |
| Essink, Brandon | Meridian Clinical Research | Omaha, NE, USA |
| Falcone, Robert | Amici Clinical Research | Raritan, NJ, USA |
| Falsey, Ann | University of Rochester/Rochester General Hospital | Rochester, NY, USA |
| Farrington, Cecil | PMG Research of Salisbury | Salisbury, NC, USA |

| | | |
|---|---|---|
| Finberg, Robert | University of Massachusetts Medical School | Worcester, MA, USA |
| Finn, Daniel | Kentucky Pediatric/Adult Research | Bardstown, KY, USA |
| Fitz-Patrick, David | East-West Medical Research Institute | Honolulu, HI, USA |
| Fortmann, Stephen | Kaiser Permanente Center for Health Research | Portland, OR, USA |
| Fouche, Leon | Tambotie Medical Centre | Thabazimbi, South Africa |
| Fragoso, Veronica | Texas Center for Drug Development | Houston, TX, USA |
| Frenck, Robert | Cincinnati Children’s Hospital | Cincinnati, OH, USA |
| Fried, David | Omega Medical Research | Warwick, RI, USA |
| Fuller, Gregory | Ventavia Research Group | Keller, TX, USA |
| Fussell, Suzanne | Long Beach Clinical Trials Services | Long Beach, CA, USA |
| Garcia-Diaz, Julia | Ochsner Health | New Orleans, LA, USA |
| Gentry, Andrew | Bozeman Health Clinical Research | Bozeman, MT, USA |
| Glover, Richard | Alliance for Multispecialty Research | Newton, KS, USA |
| Greenbaum, Carla | Benaroya Research Institute at Virginia Mason | Seattle, WA, USA |
| Grubb, Stephen | Main Street Physicians | Little River, SC, USA |
| Hammitt, Laura | Johns Hopkins Center for American Indian Health | Whiteriver, AZ, USA |
| | Johns Hopkins Center for American Indian Health | Chinle, AZ, USA |
| | Johns Hopkins Center for American Indian Health | Shiprock, NM, USA |
| | Johns Hopkins Center for American Indian Health | Gallup, NM, USA |
| Harper, Charles | Meridian Clinical Research | Norfolk, NE, USA |
| Harper, Wayne | Wake Research | Raleigh, NC, USA |
| Hartman, Aaron | Virginia Research Center | Midlothian, VA, USA |
| Heller, Robert | Bayview Research Group | Valley Village, CA, USA |
| Hendrix, Ernest | North Alabama Research Center | Athens, AL, USA |
| Herrington, Darrell | Benchmark Research | San Angelo, TX, USA |
| Jennings, Timothy | Clinical Research Professionals | Chesterfield, MO, USA |
| Karabay, Oğuz | Sakarya University Faculty of Medicine | Sakarya, Turkey |
| Kaster, Steven | Confluence Health/Wenatchee Valley Hospital & Clinics | Wenatchee, WA, USA |
| Katzman, Steven | Michigan Center for Medical Research | Farmington Hills, MI, USA |
| Kingsley, Jeffrey | Columbus Regional Research Institute | Columbus, GA, USA |
| Klein, Nicola | Kaiser Permanente, Northern California | Santa Clara, CA, USA |
| | | Sacramento, CA, USA |
| Klein, Tracy | Heartland Research Associates | Wichita, KS, USA |
| Koch, Mark | Ventavia Research Group | Fort Worth, TX, USA |
| Köksal, Iftahar | Acibadem University Hospital Atakent | Istanbul, Turkey |
| Koren, Michael | Jacksonville Center for Clinical Research | Jacksonville, FL, USA |
| Kutner, Mark | Suncoast Research Group | Miami, FL, USA |
| Lee, Marcus | Trinity Clinical Research | Tullahoma, TN, USA |
| Leibowitz, Mark | National Research Institute | Huntingdon Park, CA, USA |
| Levin, Michael | Clinical Research Center of Nevada | Las Vegas, NV, USA |
| Libster, Romina | Fundacion INFANT | Buenos Aires, Argentina |
| Lillestol, Michael | Lillestol Research | Fargo, ND, USA |
| Lucasti, Christopher | South Jersey Infectious Disease | Somers Point, NJ, USA |
| Luttermann, Matthias | Studienzentrum Brinkum Dr. Lars Pohlmeier und Torsten Drescher | Stuhr, Germany |
| Manning, Mary Beth | Velocity Clinical Research | Cleveland, OH, USA |
| Martin, Earl | Martin Diagnostic Clinic | Tomball, TX, USA |
| Matherne, Paul | MedPharmics | Gulfport, MS, USA |

| | | |
|---|---|---|
| McMurray, James | Medical Affiliated Research Center (MARC) | Huntsville, AL, USA |
| Mert, Ali | Medipol University Hospital | Istanbul, Turkey |
| Middleton, Randle | Optimal Research | Huntsville, AL, USA |
| Mitha, Essack | Newtown Clinical Research | Johannesburg, South Africa |
| Morawski, Emily | Holston Medical Group | Kingsport, TN, USA |
| Moreira, Edson | Associação Obras Sociais Irmã Dulce and Oswaldo Cruz Foundation | Bahia, Brazil |
| Murray, Alexander | PharmQuest | Greensboro, NC, USA |
| Mussaji, Murtaza | LinQ Research | Houston, TX, USA |
| Musungaie, Dany | Jongaie Research | Pretoria, South Africa |
| Nell, Haylene | Tiervlei Trial Centre, Karl Bremer Hospital | Cape Town, South Africa |
| Odekirk, Larry | Lynn Institute of Denver | Denver, CO, USA |
| Ogbuagu, Onyema | Yale University School of Medicine | New Haven, CT, USA |
| Paolino, Kristopher | State University of New York, Upstate Medical University | Syracuse, NY, USA |
| Patel, Suchet | Regional Clinical Research | Endwell, NY, USA |
| Peterson, James | J. Lewis Research | Salt Lake City, UT, USA |
| Pickrell, Paul | Tekton Research | Austin, TX, USA |
| Polack, Fernando | Fundacion INFANT | Buenos Aires, Argentina |
| Poretz, Donald | Clinical Alliance for Research and Education | Annandale, VA, USA |
| Raad, George | PMG Research of Charlotte | Charlotte, NC, USA |
| Randall, William | PriMed Clinical Research | Dayton, OH, USA |
| Rankin, Bruce | Accel Research Sites - DeLand Clinical Research Unit | DeLand, FL, USA |
| Reynolds, Steven | Collaborative Neuroscience Network | Long Beach, CA, USA |
| Riesenberg, Robert | Atlanta Center for Medical Research | Atlanta, GA, USA |
| Rodriguez, Hector | Acevedo Clinical Research Associates | Miami, FL, USA |
| Rosen, Jeffrey | Clinical Research of South Florida | Coral Gables, FL, USA |
| Rubino, John | Raleigh Medical Group, PA | Raleigh, NC, USA |
| Rupp, Richard | University of Texas | Galveston, TX, USA |
| Saiger, Salma | Research Across America | Mesquite, TX, USA |
| Salata, Robert | University Hospitals Cleveland Medical Center | Cleveland, OH, USA |
| Saleh, Jamshid | Northern California Neurological Surgery | Redding, CA, USA |
| Schaefer, Axel | Medizentrum Essen Borbeck | Essen, Germany |
| Schear, Martin | Dayton Clinical Research | Dayton, OH, USA |
| Schultz, Armin | CRS Clinical Research Services Mannheim | Mannheim, Germany |
| Schwartz, Howard | Research Centers of America | Hollywood, FL, USA |
| Segall, Nathan | Clinical Research Atlanta | Stockbridge, GA, USA |
| Seger, William | HealthFirst Medical Group | Fort Worth, TX, USA |
| Senders, Shelly | Senders Pediatrics | South Euclid, OH, USA |
| Sharp, Stephan | Clinical Research Associates | Nashville, TN, USA |
| Shoffner, Sylvia | PMG Research of Cary | Cary, NC, USA |
| Simsek Yavuz, Serap | Istanbul Faculty of Medicine, Istanbul University | Istanbul, Turkey |
| Sligh, Teresa | Providence Clinical Research | North Hollywood, CA, USA |
| Smith, William | New Orleans Center for Clinical Research | New Orleans, LA, USA |
| Stacey, Helen | Diablo Clinical Research | Walnut Creek, CA, USA |
| Stephens, Michael | Fleming Island Center for Clinical Research | Fleming Island, FL, USA |
| Studdard, Harry | Coastal Clinical Research | Mobile, AL, USA |
| Tabak, Fehmi | Istanbul University Cerrahpaşa Medical School | Istanbul, Turkey |
| Talaat, Kawsar | Johns Hopkins School of Medicine | Baltimore, MD, USA |

| | | |
|---|---|---|
| Thomas, Stephen | State University of New York, Upstate Medical University | Syracuse, NY, USA |
| Towner, William | Kaiser Permanente | Los Angeles, CA, USA |
| Tran, Van | Ventavia Research Group | Houston, TX, USA |
| Ünal, Serhat | Hacettepe University | Ankara, Turkey |
| Usdan, Lisa | CNS Healthcare | Memphis, TN, USA |
| Vanchiere, John | Louisiana State University Health Shreveport | Shreveport, LA, USA |
| Varano, Susann | Clinical Research Consulting | Milford, CT, USA |
| Wadsworth, L. Tyler | Sundance Clinical Research | St Louis, MO, USA |
| Walsh, Edward | University of Rochester/Rochester General Hospital | Rochester, NY, USA |
| Walter, Emmanuel | Duke Human Vaccine Institute | Durham, NC, USA |
| Wappner, Diego | Fundacion INFANT | Buenos Aires, Argentina |
| Whiles, Rick | Holston Medical Group | Bristol, TN, USA |
| Williams, Hayes | Achieve Clinical Research | Birmingham, AL, USA |
| Wilson, Jonathan | Piedmont Medical Research of Winston-Salem | Winston-Salem, NC, USA |
| Winkle, Peter | Anaheim Clinical Trials | Anaheim, CA, USA |
| Winokur, Patricia | University of Iowa | Iowa City, IA, USA |
| Wolf, Thomas | Prairie Fields Family Medicine | Fremont, NE, USA |
| Yozviak, Joseph | Lehigh Valley Hospital Cedar Crest | Allentown, PA, USA |
| Zerbini, Cristiano | Centro Paulista de Investigação Clínica - CEPIC | São Paulo, Brazil |

### Ethical Conduct of the Study

The trial was conducted in accordance with the protocol, the ethical principles derived from international guidelines including the International Council for Harmonisation Good Clinical Practice Guidelines, the Declaration of Helsinki and Council for International Organizations of Medical Sciences International Ethical Guidelines, and applicable laws and regulations (including applicable privacy laws). An independent data monitoring committee reviewed efficacy and unblinded safety data. Institutional Review Board or Ethics Committee approval was obtained for each site prior to enrollment of any study participant.

The list of Institutional Review Board Committees is summarized at the end of this Supplementary Appendix.

### Study Responsibilities

Pfizer was responsible for the design, study conduct, data collection, data analysis, data interpretation, and writing of this manuscript. Both Pfizer and BioNTech manufactured clinical trial material. BioNTech was the sponsor of the study and contributed to data interpretation and writing of the manuscript. All study data were available to all authors who vouch for its accuracy and adherence of the study to the protocol.

### Testing for SARS-CoV-2 Virus and Antibodies

Testing for SARS-CoV-2 virus was conducted using the Cepheid Xpert Xpress SARS-CoV-2 RT-PCR test. Testing for SARS-CoV-2 antibodies was conducted using the Roche Elecsys® Anti-SARS-CoV-2 antibody test.

### Determination of SARS-CoV-2 Lineage

For determination of SARS-CoV-2 lineage, nucleic acid extraction of midturbinate swab specimens was performed using the MagMAX™ Viral/Pathogen Ultra Nucleic Acid Isolation Kit processed on a KingFisher™ Presto.

SARS-CoV-2 viral genome sequencing was performed using the Ion Torrent and Illumina NextSeq platforms. For the Ion Torrent sequencing platform, the Ion AmpliSeq™ SARS-CoV-2 Research Panel was used, which consists of 2 primer pools targeting a total of 237 PCR amplicons specific to SARS-CoV-2 and 5 human expression controls in each pool. Oligonucleotide primers based on available SARS-CoV-2 nucleotide sequences direct the amplification of the viral genome with amplicon lengths of 125–275 bp. The panel provides >99% coverage of the SARS-CoV-2 genome (~30 kb). To determine the optimal number of target amplification cycles, SARS-CoV-2 viral RNA content in the nucleic acid purified from the midturbinate specimens was quantified using the TaqMan™ 2019-nCoV Assay Kit v1, the TaqMan™ 2019-nCoV Control Kit v1, and TaqPath™ 1-Step RT-qPCR Master Mix, CG. cDNA was synthesized with the SuperScript VILO cDNA synthesis kit. Libraries were prepared using the Ion AmpliSeq™ Library Kit plus the Ion AmpliSeq™ SARS-CoV-2 Research Panel according to the manufacturer's instructions (ThermoFisher. Ion AmpliSeq™ Library Kit Plus USER GUIDE. Publication MAN0017003 version C.0.). Libraries underwent template preparation with Ion Chef according to the manufacturer's instructions. Prepared templates were loaded onto an Ion 530 chip for semiconductor sequencing on the Ion GeneStudio™ S5 plus sequencer according to the manufacturer's instructions. Raw sequencing reads generated by the Ion Torrent sequencer were quality and adaptor trimmed by Ion Torrent Suite and the resulting reads were then mapped to the complete genome of the SARS-CoV-2 Wuhan-Hu-1 isolate (GenBank accession number MN908947.3) using TMAP 5.14.0. Variant calling was carried out with the Torrent Variant Caller using the BAM file from the mapping of the cleaned sequence reads onto the reference sequence of SARS-CoV-2.

SARS-CoV-2 viral genome sequencing performed using the Illumina NextSeq platform used the AmpliSeq for Illumina SARS-CoV-2 panel of PCR primers to enrich for SARS-CoV-2 in the biological specimen. This was a 2-pool design, containing a total of 237 SARS-CoV-2 amplicon/primer pairs plus 5 human expression controls in each pool. Oligonucleotide primers based on available SARS-CoV-2 nucleotide sequences directed the amplification of overlapping amplicons with lengths of 125–275 bp that cover >99% of the viral genome. Nucleic acid extracted from the midturbinate specimens was digested initially with DNase (Invitrogen TURBO DNA-free™ Kit, AM1907), and RNA was purified using MagMAX™ beads before cDNA synthesis. Synthesis of cDNA using random sequence primers and downstream steps were as described by the manufacturer. SARS-CoV-2 amplicons were generated from the cDNA, followed by ligation of Universal Next Generation Sequencing Adaptors to the ends of the amplicons. Amplicon libraries were purified with magnetic beads and loaded onto a flow cell for sequence determination using the Illumina NextSeq instrument, according to the manufacturer's instructions. Sequences with ≥30-fold coverage across the entire spike gene were advanced for viral lineage assignment. Single nucleotide variants were called using the "Low Frequency Variant Detection" function with the cut-off for sequence heterogeneity set at >10%.

SARS-CoV-2 lineage assignment was based on Pangolin 2.0 software, which runs a multinomial logistic regression model trained against lineage assignments based on isolate data from the Global Initiative on Sharing All Influenza Data (GISAID), a global science initiative established in 2008 that provides open-access to genomics data of influenza virus and SARS-CoV-2.

**Definitions of Confirmed and Severe COVID-19 Cases**

The definition of SARS-CoV-2-related cases was the presence of ≥1 of the following symptoms and SARS-CoV-2-NAAT positivity during or within 4 days before or after the symptomatic period: fever, new or increased cough, new or increased shortness of breath, chills, new or increased muscle pain, new loss of taste or smell, sore throat, diarrhea, and/or vomiting. The onset date of the case was the date that symptoms were first experienced by the participant. If new symptoms were reported ≤4 days after resolution of all previous symptoms, they were considered part of a single illness.

Confirmed severe COVID-19 required confirmation of COVID-19 and the presence of ≥1 of the following: clinical signs at rest indicative of severe systemic illness (respiratory rate ≥30 breaths per minute, heart rate ≥125 beats per minute, $SpO_2$ ≤93% on room air at sea level, or $PaO_2/FiO_2$ <300 mmHg); respiratory failure (defined as needing high-flow oxygen, non-invasive ventilation, mechanical ventilation, or extracorporeal membrane oxygenation); evidence of shock (systolic blood pressure <90 mmHg, diastolic blood pressure <60 mmHg, or requiring vasopressors); significant acute renal, hepatic, or neurologic dysfunction; intensive care unit admission; and/or death (https://www.fda.gov/media/137926/download).

| Figure/Table Number | Figure/Table Title | Population(s)/Sample Size | Explanation |
|---|---|---|---|
| Figure 1 | Disposition of Participants | All enrolled safety population ≥16 years of age<br>N=44,165 | Per protocol |
| Figure 2 | Local Reactions and Systemic Events Reported within 7 Days after Receipt of BNT162b2 or Placebo by Baseline SARS-CoV-2 Status | Reactogenicity subset of participants ≥16 years of age (ie, participants who used an electronic diary for reporting local reactions and systemic events)<br>N=9839 | Per protocol |
| Figure 3 | Efficacy of BNT162b2 against COVID-19 Occurrence after Dose 1 During the Placebo-controlled Follow-up Period | N=46,077 (all available) | All randomized participants ≥12 years of age |
| Table 1 | Demographics | Participants ≥16 years of age<br>N=44,047 | Includes HIV-infected individuals |
| Table 2 | Vaccine Efficacy against COVID-19 from 7 Days after Dose 2 During the Blinded Placebo Controlled Follow-up Period (Evaluable Efficacy Population, ≥12 Years Old) | a. Efficacy endpoint including individuals **without** evidence of prior infection (N=42,094)<br><br>b. Efficacy endpoint including individuals **with and those without** evidence of prior infection (N=44,486) | Evaluable population:<br>• received 2 vaccinations as randomized<br>• no major protocol deviations<br>Excludes HIV+ participants |
| Table 3 | Vaccine Efficacy Overall and by Subgroup in Participants Without Evidence of Infection Prior to 7 Days After Dose 2 During the Blinded Placebo Controlled Follow-up Period | N=42,094 (same as efficacy endpoint in Table 2, participants ≥12 years of age) | |
| Table S2 | Baseline Comorbidities in Participants ≥16 Years of Age | N=44,047 | Includes HIV-infected individuals |
| Table S3 | Participants Reporting at Least 1 Adverse Event From Dose 1 to 1 Month After Dose 2 During the Blinded Follow-up Period | Participants ≥16 years of age<br>N=43,847 | Vaccinated minus 200 HIV-infected participants |
| Table S4 | Causes of Death from Dose 1 to Unblinding (Safety Population, ≥16 Years Old) | Participants ≥16 years of age<br>N=43,847 | Vaccinated minus 200 HIV-infected participants |
| Table S5 | Vaccine Efficacy Overall and by Subgroup after Dose 1 During the Blinded Placebo Controlled Follow-up Period (All-Available Population) | N=46,077 (all available) | |
| Table S6 | Vaccine Efficacy against Severe COVID-19 Occurrence after Dose 1 (All-Available Population) | N=46,077 (all available, participants ≥12 years of age) | |
| Table S7 | Vaccine Efficacy from 7 Days after Dose 2 by Underlying Comorbidities (Risk Status) among Participants without Evidence of Infection Prior to 7 Days after Dose 2 (Evaluable Efficacy Population) | N=42,094 (same as efficacy endpoint in Table 2, participants ≥12 years of age) | |

**Table S1 | Explanation of the Changes in Denominator Numbers in Various Analyses.**

| Charlson Comorbidity Index Category | BNT162b2 (N[a]=22,026) | Placebo (N[a]=22,021) | Total (N[a]=44,047) |
|---|---|---|---|
| | n[b] (%) | n[b] (%) | n[b] (%) |
| Participants with any Charlson comorbidity | 4628 (21.0) | 4511 (20.5) | 9139 (20.7) |
| AIDS/HIV | 100 (0.5) | 100 (0.5) | 200 (0.5) |
| Any malignancy | 812 (3.7) | 757 (3.4) | 1569 (3.6) |
| Cerebrovascular disease | 227 (1.0) | 198 (0.9) | 425 (1.0) |
| Chronic pulmonary disease | 1783 (8.1) | 1775 (8.1) | 3558 (8.1) |
| Congestive heart failure | 109 (0.5) | 102 (0.5) | 211 (0.5) |
| Dementia | 7 (0.0) | 11 (0.0) | 18 (0.0) |
| Diabetes with chronic complication | 116 (0.5) | 130 (0.6) | 246 (0.6) |
| Diabetes without chronic complication | 1700 (7.7) | 1699 (7.7) | 3399 (7.7) |
| Hemiplegia or paraplegia | 15 (0.1) | 25 (0.1) | 40 (0.1) |
| Leukemia | 14 (0.1) | 11 (0.0) | 25 (0.1) |
| Lymphoma | 26 (0.1) | 36 (0.2) | 62 (0.1) |
| Metastatic solid tumor | 4 (0.0) | 3 (0.0) | 7 (0.0) |
| Mild liver disease | 152 (0.7) | 115 (0.5) | 267 (0.6) |
| Moderate or severe liver disease | 2 (0.0) | 3 (0.0) | 5 (0.0) |
| Myocardial infarction | 225 (1.0) | 218 (1.0) | 443 (1.0) |
| Peptic ulcer disease | 63 (0.3) | 84 (0.4) | 147 (0.3) |
| Peripheral vascular disease | 144 (0.7) | 139 (0.6) | 283 (0.6) |
| Renal disease | 140 (0.6) | 153 (0.7) | 293 (0.7) |
| Rheumatic disease | 75 (0.3) | 71 (0.3) | 146 (0.3) |

**Table S2 | Baseline Comorbidities in Participants ≥16 Years of Age.** Baseline comorbid conditions are classified according to the Charlson Comorbidity Index (Charlson M, Szatrowski TP, Peterson J, Gold J. Validation of a combined comorbidity index. J Clin Epidemiol 1994;47:1245-51.). a. N=number of participants in the specified group. This value is the denominator for the percentage calculations. b. n=number of participants with the specified characteristic. Participants with multiple occurrences within each category are counted only once. For 'Participants with any Charlson comorbidity', n=number of participants reporting ≥1 occurrence of any Charlson comorbidity.

| Adverse Event | BNT162b2 (N[a]=21,926) n[b] (%) | Placebo (N[a]=21,921) n[b] (%) |
|---|---|---|
| Any event | 6617 (30.2) | 3048 (13.9) |
| Related[c] | 5241 (23.9) | 1311 (6.0) |
| Severe | 262 (1.2) | 150 (0.7) |
| Life-threatening | 21 (0.1) | 26 (0.1) |
| Any serious adverse event | 127 (0.6) | 116 (0.5) |
| Related[c,d] | 3 (0.0) | 0 |
| Severe | 71 (0.3) | 66 (0.3) |
| Life-threatening | 21 (0.1) | 26 (0.1) |
| Any adverse event leading to withdrawal | 32 (0.1) | 36 (0.2) |
| Related[c] | 13 (0.1) | 11 (0.1) |
| Severe | 10 (0.0) | 10 (0.0) |
| Life-threatening | 3 (0.0) | 7 (0.0) |
| Death | 3 (0.0) | 5 (0.0) |

**Table S3 | Participants Reporting at Least 1 Adverse Event from Dose 1 to 1 Month After Dose 2 During the Blinded Follow-up Period.** The population included all ≥16-year-old participants who received ≥1 dose of vaccine irrespective of follow-up time. a. N=number of participants in the specified group. This value is the denominator for the percentage calculations. b. n=Number of participants reporting ≥1 occurrence of the specified event category. For 'any event', n=number of participants reporting ≥1 occurrence of any event. c. Assessed by the investigator as related to investigational product. d. Shoulder injury related to vaccine administration, right axillary lymphadenopathy, and paroxysmal ventricular arrhythmia (as previously reported). Adverse events for 12–15-year-old participants were reported previously.[11]

| Reported Cause of Death[a] | BNT162b2 (N=21,926) n | Placebo (N=21,921) n |
|---|---|---|
| Deaths | 15 | 14 |
| Acute respiratory failure | 0 | 1 |
| Aortic rupture | 0 | 1 |
| Arteriosclerosis | 2 | 0 |
| Biliary cancer metastatic | 0 | 1 |
| COVID-19 | 0 | 2 |
| COVID-19 pneumonia | 1 | 0 |
| Cardiac arrest | 4 | 1 |
| Cardiac failure congestive | 1 | 0 |
| Cardiorespiratory arrest | 1 | 1 |
| Chronic obstructive pulmonary disease | 1 | 0 |
| Death | 0 | 1 |
| Dementia | 0 | 1 |
| Emphysematous cholecystitis | 1 | 0 |
| Hemorrhagic stroke | 0 | 1 |
| Hypertensive heart disease | 1 | 0 |
| Lung cancer metastatic | 1 | 0 |
| Metastases to liver | 0 | 1 |
| Missing | 0 | 1 |
| Multiple organ dysfunction syndrome | 0 | 2 |
| Myocardial infarction | 0 | 2 |
| Overdose | 0 | 1 |
| Pneumonia | 0 | 2 |
| Sepsis | 1 | 0 |
| Septic shock | 1 | 0 |
| *Shigella* sepsis | 1 | 0 |
| Unevaluable event | 1 | 0 |

**Table S4 | Causes of Death from Dose 1 to Unblinding (Safety Population, ≥16 Years Old).** a. Multiple causes of death could be reported for each participant. There were no deaths among 12–15-year-old participants.

| First COVID-19 Occurrence after Dose 1 | BNT162b2 (N[a]=23,040) n1[b] | BNT162b2 (N[a]=23,040) Surveillance Time[c] (n2[d]) | Placebo (N[a]=23,037) n1[b] | Placebo (N[a]=23,037) Surveillance Time[c] (n2[d]) | VE (%) | (95% CI[e]) |
|---|---|---|---|---|---|---|
| Overall (≥12 years old) | 131 | 8.412 (22,505) | 1034 | 8.124 (22,434) | 87.8 | (85.3, 89.9) |
| **Efficacy endpoint by subgroup** | | | | | | |
| Select age groups (years) | | | | | | |
| 16 to 17 | 3 | 0.094 (373) | 19 | 0.090 (370) | 84.8 | (48.4, 97.1) |
| 16 to 55 | 95 | 4.845 (12,645) | 693 | 4.669 (12,626) | 86.8 | (83.6, 89.5) |
| >55 | 33 | 3.310 (8740) | 306 | 3.204 (8689) | 89.6 | (85.0, 92.9) |
| ≥65 | 12 | 1.645 (4455) | 138 | 1.596 (4437) | 91.6 | (84.8, 95.7) |
| ≥75 | 2 | 0.326 (905) | 26 | 0.310 (877) | 92.7 | (70.7, 99.2) |
| Sex | | | | | | |
| Male | 70 | 4.355 (11,560) | 500 | 4.115 (11,312) | 86.8 | (83.0, 89.9) |
| Female | 61 | 4.057 (10,945) | 534 | 4.009 (11,122) | 88.7 | (85.3, 91.5) |
| Race | | | | | | |
| White | 115 | 6.957 (18,538) | 916 | 6.719 (18,479) | 87.9 | (85.3, 90.1) |
| Black or African American | 6 | 0.783 (2042) | 53 | 0.770 (2063) | 88.9 | (74.1, 96.1) |
| American Indian or Alaska Native | 1 | 0.061 (216) | 7 | 0.055 (209) | 86.9 | (–1.6, 99.7) |
| Asian | 4 | 0.348 (995) | 26 | 0.337 (990) | 85.1 | (57.0, 96.2) |
| Native Hawaiian or other Pacific Islander | 0 | 0.021 (58) | 1 | 0.011 (32) | 100.0 | (–2000.0, 100.0) |
| Multiracial | 5 | 0.208 (565) | 25 | 0.190 (546) | 81.8 | (51.6, 94.6) |
| Not reported | 0 | 0.035 (91) | 6 | 0.042 (115) | 100.0 | (–0.7, 100.0) |
| Ethnicity | | | | | | |
| Hispanic/Latinx | 52 | 2.351 (5701) | 302 | 2.282 (5673) | 83.3 | (77.5, 87.8) |
| Non-Hispanic/non-Latinx | 78 | 6.018 (16,692) | 730 | 5.799 (16,647) | 89.7 | (87.0, 92.0) |
| Not reported | 1 | 0.043 (112) | 2 | 0.043 (114) | 49.4 | (–872.9, 99.1) |
| Country | | | | | | |
| Argentina | 32 | 1.282 (2846) | 146 | 1.269 (2840) | 78.3 | (68.0, 85.7) |
| Brazil | 14 | 0.554 (1430) | 95 | 0.520 (1420) | 86.1 | (75.6, 92.7) |
| Germany | 2 | 0.067 (246) | 1 | 0.069 (250) | –104.5 | (–11,965.9, 89.4) |
| South Africa | 0 | 0.128 (367) | 11 | 0.125 (365) | 100.0 | (61.1, 100.0) |
| Turkey | 3 | 0.048 (246) | 12 | 0.045 (244) | 76.4 | (12.4, 95.7) |
| USA | 80 | 6.333 (17,370) | 769 | 6.095 (17,315) | 90.0 | (87.4, 92.1) |
| Baseline SARS-CoV-2 status | | | | | | |
| Positive[f] | 13 | 0.250 (692) | 17 | 0.265 (736) | 19.2 | (–76.6, 63.9) |
| Positive N-binding only | 2 | 0.192 (521) | 7 | 0.198 (542) | 70.5 | (–54.7, 97.0) |

| | | | | | | |
|---|---|---|---|---|---|---|
| Positive NAAT only | 10 | 0.020 (66) | 9 | 0.020 (69) | –10.5 | (–207.3, 59.7) |
| Positive NAAT and N-binding | 1 | 0.038 (105) | 1 | 0.046 (124) | –20.5 | (–9359.2, 98.5) |
| Negative[g] | 116 | 8.101 (21,615) | 1015 | 7.804 (21,521) | 89.0 | (86.6, 91.0) |
| Unknown | 2 | 0.061 (198) | 2 | 0.055 (177) | 9.7 | (–1145.4, 93.5) |

**Table S5 | Vaccine Efficacy Overall and by Subgroup after Dose 1 During the Blinded Placebo Controlled Follow-up Period (All-Available Population).** Efficacy data are presented for participants ≥12 years old. a. N=number of participants in the specified group. b. n1=Number of participants meeting the endpoint definition. c. Total surveillance time in 1000 person-years for the given endpoint across all participants within each group at risk for the endpoint. Time period for COVID-19 case accrual is from dose 1 to the end of the surveillance period. d. n2=number of participants at risk for the endpoint. e. CI for vaccine efficacy is derived based on the Clopper and Pearson method adjusted for surveillance time. f. Positive N-binding antibody result at Visit 1, positive NAAT result at Visit 1, or medical history of COVID-19. g. Negative N-binding antibody result at Visit 1, negative NAAT result at Visit 1, and no medical history of COVID-19.

| Efficacy Endpoint<br>Subgroup | BNT162b2<br>(N[a]=23,040)<br>n1[b] | <br><br>Surveillance Time[c] (n2[d]) | Placebo<br>(N[a]=23,037)<br>n1[b] | <br><br>Surveillance Time[c] (n2[d]) | VE (%) | (95% CI[e]) |
|---|---|---|---|---|---|---|
| First severe COVID-19 occurrence after dose 1 | 1 | 8.439 (22,505) | 30 | 8.288 (22,435) | 96.7 | (80.3, 99.9) |
| After dose 1 to before dose 2 | 0 | 1.351 (22,505) | 6 | 1.360 (22,435) | 100.0 | (14.5, 100.0) |
| Dose 2 to 7 days after dose 2 | 0 | 0.425 (22,170) | 1 | 0.423 (22,070) | 100.0 | (–3783.5, 100.0) |
| ≥7 Days after dose 2 | 1 | 6.663 (22,142) | 23 | 6.505 (22,048) | 95.7 | (73.9, 99.9) |

**Table S6 | Vaccine Efficacy against Severe COVID-19 Occurrence after Dose 1 (All-Available Population).** Efficacy data are presented for participants ≥12 years old. a. N=number of participants in the specified group. b. n1=number of participants meeting the endpoint definition. c. Total surveillance time in 1000 person-years for the given endpoint across all participants within each group at risk for the endpoint. Time period for severe COVID-19 case accrual is from dose 1 to the end of the surveillance period for the overall row, and from the start to the end of the range stated for each time interval. d. n2=number of participants at risk for the endpoint. [e]. CI for vaccine efficacy is derived based on the Clopper and Pearson method adjusted for surveillance time. Severe COVID-19 as defined by the US FDA [https://www.fda.gov/media/137926/download]).

| Efficacy Endpoint<br>Subgroup | BNT162b2 ($N^a$=20,998)<br>n1[b] | <br>Surveillance Time[c] (n2[d]) | Placebo ($N^a$=21,096)<br>n1[b] | <br>Surveillance Time[c] (n2[d]) | VE (%) | (95% CI[e]) |
|---|---|---|---|---|---|---|
| **First COVID-19 occurrence from 7 days after dose 2** | | | | | | |
| Overall | 77 | 6.247 (20,712) | 850 | 6.003 (20,713) | 91.3 | (89.0, 93.2) |
| At risk[f] | | | | | | |
| Yes | 35 | 2.797 (9167) | 401 | 2.681 (9136) | 91.6 | (88.2, 94.3) |
| No | 42 | 3.450 (11,545) | 449 | 3.322 (11,577) | 91.0 | (87.6, 93.6) |
| Age group (years) and at risk | | | | | | |
| 16−64 and at risk | 29 | 2.083 (6632) | 325 | 1.993 (6629) | 91.5 | (87.5, 94.4) |
| ≥65 and at risk | 6 | 0.680 (2322) | 71 | 0.656 (2304) | 91.8 | (81.4, 97.1) |
| Obese[g] | | | | | | |
| Yes | 27 | 2.103 (6796) | 314 | 2.050 (6875) | 91.6 | (87.6, 94.6) |
| No | 50 | 4.143 (13,911) | 536 | 3.952 (13,833) | 91.1 | (88.1, 93.5) |
| Age group (years) and obese | | | | | | |
| 16−64 and obese | 24 | 1.680 (5303) | 266 | 1.624 (5344) | 91.3 | (86.7, 94.5) |
| ≥65 and obese | 3 | 0.404 (1370) | 45 | 0.410 (1426) | 93.2 | (78.9, 98.7) |

**Table S7 | Vaccine Efficacy from 7 Days after Dose 2 by Underlying Comorbidities (Risk Status) among Participants Without Evidence of Infection Prior to 7 Days after Dose 2 (Evaluable Efficacy Population).** Efficacy data are presented for participants ≥12 years old. a. N=number of participants in the specified group. b. n1=number of participants meeting the endpoint definition. c. Total surveillance time in 1000 person-years for the given endpoint across all participants within each group at risk for the endpoint. Time period for COVID-19 case accrual is from 7 days after dose 2 to the end of the surveillance period. d. n2=number of participants at risk for the endpoint. e. CI for vaccine efficacy is derived based on the Clopper and Pearson method adjusted for surveillance time. f. Includes participants who had ≥1 Charlson Comorbidity Index (CMI) category or obesity (body mass index [BMI] ≥30 $kg/m^2$ [≥16 years old] or BMI ≥95th percentile [12–15 years old]). g. Participants who had BMI ≥30 $kg/m^2$ (≥16 years old) or BMI ≥95th percentile (12–15 years old; refer to the CDC growth charts at https://www.cdc.gov/growthcharts/html_charts/bmiagerev.htm).

### Independent Ethics Committee or Institutional Review Board Address(es)

| |
|---|
| Comité Institucional de Revisión de Ensayos Clínicos (C.I.R.E.C.) del Hospital Militar Central "Cirujano Mayor Dr Cosme Argerich"<br>Av. Luis María Campos 726, Edificio PACE Piso 5<br>CABA, 1426<br>ARGENTINA |
| CONEP (Comissao Nacional de Etica em Pesquisa)<br>SRTV 701, Via W 5 Norte, lote D, Edificio PO 700, 3º andar - Asa Norte<br>Brasilia, DF 70719-040<br>BRAZIL |
| Comite de Etica em Pesquisa da Faculdade de Medicina do ABC\Fundacao do ABC-- FMABC<br>Avenida Lauro Gomes, 2000 - Vila Sacadura Cabral<br>Santo Andre/SP, 09060-870<br>BRAZIL |
| Comite de Etica em Pesquisa do Hospital Santo Antonio /Obras Sociais Irma Dulce<br>Avenida Luiz Tarquínio, snº, portao 9, 1º andar, sala 1, Roma<br>Salvador, BA 40414-120<br>BRAZIL |
| CONEP (Comissão Nacional de Ética em Pesquisa)<br>SRTV 701, Via W 5 Norte, lote D, Edifício PO 700, 3° andar -Asa Norte<br>Brasília/DF, 70719-040<br>BRAZIL |
| Landesaerztekammer Baden-Wuerttemberg<br>Liebknechtstr. 33<br>Stuttgart, 70565<br>GERMANY |
| Landesaerztekammer Baden-Wuerttemberg<br>Liebknechtstr. 33<br>Stuttgart, 70565<br>GERMANY |
| Landesaerztekammer Baden-Wuerttemberg<br>Liebknechtstr. 33<br>Stuttgart, 70565<br>GERMANY |
| Landesaerztekammer Baden-Wuerttemberg<br>Liebknechtstr. 33<br>Stuttgart, 70565<br>GERMANY |
| Landesaerztekammer Baden-Wuerttemberg<br>Liebknechtstr. 33<br>Stuttgart, 70565<br>GERMANY |
| Landesaerztekammer Baden-Wuerttemberg<br>Liebknechtstr. 33<br>Stuttgart, 70565<br>GERMANY |
| Pharma Ethics Independent Research Ethics committee<br>123 Amcor Road, Lyttelton Manor<br>Centurion, 0157<br>SOUTH AFRICA |
| Pharma-Ethics (Pty) Ltd<br>123 Amkor Road, Lyttelton Manor<br>Centurion, 0157<br>SOUTH AFRICA |

| |
|---|
| Pharma-Ethics (Pty) Ltd<br>123 Amkor Road, Lyttelton Manor<br>Centurion, 0157<br>SOUTH AFRICA |
| Pharma-Ethics (Pty) Ltd<br>123 Amkor Road, Lyttelton Manor<br>Centurion, 0157<br>SOUTH AFRICA |
| Kocaeli Üniversitesi Klinik Arastirmalar Etik Kurulu<br>Kocaeli Üniversitesi Tip Fakültesi Klinik, Arastirmalar Birimi Umuttepe Yerleskesi<br>Kocaeli<br>TURKEY |
| Kocaeli Üniversitesi Klinik Arastirmalar Etik Kurulu<br>Kocaeli Üniversitesi Tip Fakültesi Klinik, Arastirmalar Birimi Umuttepe Yerleskesi<br>Kocaeli<br>TURKEY |
| Kocaeli Üniversitesi Klinik Arastirmalar Etik Kurulu<br>Kocaeli Üniversitesi Tip Fakültesi Klinik, Arastirmalar Birimi Umuttepe Yerleskesi<br>Kocaeli<br>TURKEY |
| Kocaeli Üniversitesi Klinik Arastirmalar Etik Kurulu<br>Kocaeli Üniversitesi Tip Fakültesi Klinik, Arastirmalar Birimi Umuttepe Yerleskesi<br>Kocaeli<br>TURKEY |
| Kocaeli Üniversitesi Klinik Arastirmalar Etik Kurulu<br>Kocaeli Üniversitesi Tip Fakültesi Klinik, Arastirmalar Birimi Umuttepe Yerleskesi<br>Kocaeli<br>TURKEY |
| Kocaeli Üniversitesi Klinik Arastirmalar Etik Kurulu<br>Kocaeli Üniversitesi Tip Fakültesi Klinik, Arastirmalar Birimi Umuttepe Yerleskesi<br>Kocaeli<br>TURKEY |
| Kocaeli Üniversitesi Klinik Arastirmalar Etik Kurulu<br>Kocaeli Üniversitesi Tip Fakültesi Klinik, Arastirmalar Birimi Umuttepe Yerleskesi<br>Kocaeli<br>TURKEY |
| Kocaeli Üniversitesi Klinik Arastirmalar Etik Kurulu<br>Kocaeli Üniversitesi Tip Fakültesi Klinik, Arastirmalar Birimi Umuttepe Yerleskesi<br>Kocaeli<br>TURKEY |
| Kocaeli Üniversitesi Klinik Arastirmalar Etik Kurulu<br>Kocaeli Üniversitesi Tip Fakültesi Klinik, Arastirmalar Birimi Umuttepe Yerleskesi<br>Kocaeli<br>TURKEY |
| NYU Langone Grossman School of Medicine IRB<br>One Park Ave, 6th Fl<br>New York, NY 10016<br>UNITED STATES |
| Western Institutional Review Board<br>1019 39th Ave., SE, Ste 120<br>Puyallup, WA 98374<br>UNITED STATES |
| Western Institutional Review Board<br>1019 39th Ave SE, Ste 120<br>Puyallup, WA 98374 |

| |
|---|
| UNITED STATES |
| Copernicus Group Institutional Review Board<br>5000 CentreGreen Way, Ste 200<br>Cary, NC 27513<br>UNITED STATES |
| Copernicus Group Institutional Review Board<br>5000 CentreGreen Way, Ste 200<br>Cary, NC 27513<br>UNITED STATES |
| Cincinnati Children's Hospital Medical Center IRB<br>3333 Burnet Ave, MLC 5020<br>Cincinnati, OH 45229<br>UNITED STATES |
| Copernicus Group Institutional Review Board<br>5000 CentreGreen Way, Ste 200<br>Cary, NC 27513<br>UNITED STATES |
| Copernicus Group Institutional Review Board<br>5000 CentreGreen Way, Ste 200<br>Cary, NC 27513<br>UNITED STATES |
| Copernicus Group Institutional Review Board<br>5000 CentreGreen Way, Ste 200<br>Cary, NC 27513<br>UNITED STATES |
| Copernicus Group Institutional Review Board<br>5000 CentreGreen Way, Ste 200<br>Cary, NC 27513<br>UNITED STATES |
| Copernicus Group IRB<br>5000 Centregreen Way, Ste 200<br>Cary, NORTH CAROLINA 27513<br>UNITED STATES |
| WCG IRB<br>1019 39th Ave Se, Ste 120<br>Puyallup, WASHINGTON 98374<br>UNITED STATES |
| Copernicus Group Institutional Review Board<br>5000 CentreGreen Way, Ste 200<br>Cary, NC 27513<br>UNITED STATES |
| Copernicus Group Institutional Review Board<br>5000 CentreGreen Way, Ste 200<br>Cary, NC 27513<br>UNITED STATES |
| Copernicus Group IRB<br>5000 CentreGreen Way, Ste 200<br>Cary, NORTH CAROLINA 27513<br>UNITED STATES |
| Copernicus Group IRB<br>5000 Centregreen Way, Ste 200<br>Cary, NORTH CAROLINA 27513<br>UNITED STATES |
| Copernicus Group Institutional Review Board<br>5000 CentreGreen Way, Ste 200 |

| |
|---|
| Cary, NC 27513<br>UNITED STATES |
| Copernicus Group Institutional Review Board<br>5000 CentreGreen Way, Ste 200<br>Cary, NC 27513<br>UNITED STATES |
| Copernicus Group Institutional Review Board<br>5000 CentreGreen Way, Ste 200<br>Cary, NC 27513<br>UNITED STATES |
| Copernicus Group IRB<br>5000 Centregreen Way, Ste 200<br>Cary, NORTH CAROLINA 27513<br>UNITED STATES |
| Copernicus Group Institutional Review Board<br>5000 CentreGreen Way, Ste 200<br>Cary, NC 27513<br>UNITED STATES |
| Copernicus Group Institutional Review Board<br>5000 CentreGreen Way, Ste 200<br>Cary, NC 27513<br>UNITED STATES |
| Copernicus Group IRB<br>5000 Centregreen Way, Ste 200<br>Cary, NORTH CAROLINA 27513<br>UNITED STATES |
| Copernicus Group Institutional Review Board<br>5000 CentreGreen Way, Ste 200<br>Cary, NC 27513<br>UNITED STATES |
| Copernicus Group Institutional Review Board<br>5000 CentreGreen Way, Ste 200<br>Cary, NC 27513<br>UNITED STATES |
| Copernicus Group IRB<br>5000 Centregreen Way, Ste 200<br>Cary, NORTH CAROLINA 27513<br>UNITED STATES |
| Copernicus Group Institutional Review Board<br>5000 CentreGreen Way, Ste 200<br>Cary, NC 27513<br>UNITED STATES |
| Copernicus Group IRB<br>5000 Centregreen Way, Ste 200<br>Cary, NORTH CAROLINA 27513<br>UNITED STATES |
| Copernicus Group Institutional Review Board<br>5000 CentreGreen Way, Ste 200<br>Cary, NC 27513<br>UNITED STATES |
| Copernicus Group Institutional Review Board<br>5000 CentreGreen Way, Ste 200<br>Cary, NC 27513<br>UNITED STATES |
| Copernicus Group Institutional Review Board |

| |
|---|
| 5000 CentreGreen Way, Ste 200<br>Cary, NC 27513<br>UNITED STATES |
| Copernicus Group Institutional Review Board<br>5000 CentreGreen Way, Ste 200<br>Cary, NC 27513<br>UNITED STATES |
| Copernicus Group IRB<br>5000 Centregreen Way, Ste 200<br>Cary, NORTH CAROLINA 27513<br>UNITED STATES |
| Copernicus Group Institutional Review Board<br>5000 Centregreen Way, Ste 200<br>Cary, NORTH CAROLINA 27513<br>UNITED STATES |
| Copernicus Group IRB<br>5000 Centregreen Way, Ste 200<br>Cary, NORTH CAROLINA 27513<br>UNITED STATES |
| Copernicus Group Institutional Review Board<br>5000 CentreGreen Way, Ste 200<br>Cary, NC 27513<br>UNITED STATES |
| Copernicus Group Institutional Review Board<br>5000 CentreGreen Way, Ste 200<br>Cary, NC 27513<br>UNITED STATES |
| Copernicus Group Institutional Review Board<br>5000 CentreGreen Way, Ste 200<br>Cary, NC 27513<br>UNITED STATES |
| Copernicus Group Institutional Review Board<br>5000 CentreGreen Way, Ste 200<br>Cary, NC 27513<br>UNITED STATES |
| Copernicus Group Institutional Review Board<br>5000 CentreGreen Way, Ste 200<br>Cary, NC 27513<br>UNITED STATES |
| Copernicus Group Institutional Review Board<br>5000 CentreGreen Way, Ste 200<br>Cary, NC 27513<br>UNITED STATES |
| Copernicus Group Institutional Review Board<br>5000 CentreGreen Way, Ste 200<br>Cary, NC 27513<br>UNITED STATES |
| Copernicus Group IRB<br>5000 Centregreen Way, Ste 200<br>Cary, NORTH CAROLINA 27513<br>UNITED STATES |
| Copernicus Group Institutional Review Board<br>5000 CentreGreen Way, Ste 200<br>Cary, NC 27513<br>UNITED STATES |

| |
|---|
| Copernicus Group Institutional Review Board<br>5000 CentreGreen Way, Ste 200<br>Cary, NC 27513<br>UNITED STATES |
| Copernicus Group Institutional Review Board<br>5000 CentreGreen Way, Ste 200<br>Cary, NC 27513<br>UNITED STATES |
| Copernicus Group Institutional Review Board<br>5000 CentreGreen Way, Ste 200<br>Cary, NC 27513<br>UNITED STATES |
| Copernicus Group IRB<br>5000 CentreGreen Way, Ste 200<br>Cary, NC 27513<br>UNITED STATES |
| Copernicus Group Institutional Review Board<br>5000 CentreGreen Way, Ste 200<br>Cary, NC 27513<br>UNITED STATES |
| Copernicus Group Institutional Review Board<br>5000 CentreGreen Way, Ste 200<br>Cary, NC 27513<br>UNITED STATES |
| Copernicus Group IRB<br>5000 Centregreen Way, Ste 200<br>Cary, NORTH CAROLINA 27513<br>UNITED STATES |
| Copernicus Group IRB<br>5000 Centregreen Way, Ste 200<br>Cary, NORTH CAROLINA 27513<br>UNITED STATES |
| Copernicus Group IRB<br>5000 Centregreen Way, Ste 200<br>Cary, NORTH CAROLINA 27513<br>UNITED STATES |
| Copernicus Group Institutional Review Board<br>5000 CentreGreen Way, Ste 200<br>Cary, NC 27513<br>UNITED STATES |
| Copernicus Group IRB<br>5000 Centregreen Way, Ste 200<br>Cary, NORTH CAROLINA 27513<br>UNITED STATES |
| Copernicus Group Institutional Review Board<br>5000 CentreGreen Way, Ste 200<br>Cary, NC 27513<br>UNITED STATES |
| Copernicus Group Institutional Review Board<br>5000 CentreGreen Way, Ste 200<br>Cary, NC 27513<br>UNITED STATES |
| Copernicus Group IRB<br>5000 CentreGreen Way, Ste 200<br>Cary, NC 27513 |

| |
|---|
| UNITED STATES |
| Copernicus Group Institutional Review Board<br>5000 CentreGreen Way, Ste 200<br>Cary, NC 27513<br>UNITED STATES |
| Copernicus Group Institutional Review Board<br>5000 Centregreen Way, Ste 200<br>Cary, NORTH CAROLINA 27513<br>UNITED STATES |
| Copernicus Group Institutional Review Board<br>5000 CentreGreen Way, Ste 200<br>Cary, NC 27513<br>UNITED STATES |
| Copernicus Group Institutional Review Board<br>5000 CentreGreen Way, Ste 200<br>Cary, NC 27513<br>UNITED STATES |
| Copernicus Group IRB<br>5000 CentreGreen Way, Ste 200<br>Cary, NORTH CAROLINA 27513<br>UNITED STATES |
| WCG IRB<br>1019 39th Ave Se, Ste 120<br>Puyallup, WASHINGTON 98374<br>UNITED STATES |
| Copernicus Group Institutional Review Board<br>5000 CentreGreen Way, Ste 200<br>Cary, NC 27513<br>UNITED STATES |
| Copernicus Group Institutional Review Board<br>5000 CentreGreen Way, Ste 200<br>Cary, NC 27513<br>UNITED STATES |
| Copernicus Group Institutional Review Board<br>5000 CentreGreen Way, Ste 200<br>Cary, NC 27513<br>UNITED STATES |
| Copernicus Group Institutional Review Board<br>5000 CentreGreen Way, Ste 200<br>Cary, NC 27513<br>UNITED STATES |
| Copernicus Group Institutional Review Board<br>5000 CentreGreen Way, Ste 200<br>Cary, NC 27513<br>UNITED STATES |
| Copernicus Group Institutional Review Board<br>5000 Centregreen Way, Ste 200<br>Cary, NORTH CAROLINA 27513<br>UNITED STATES |
| Copernicus Group Institutional Review Board<br>5000 CentreGreen Way, Ste 200<br>Cary, NC 27513<br>UNITED STATES |
| Copernicus Group Institutional Review Board<br>5000 CentreGreen Way, Ste 200 |

| |
|---|
| Cary, NC 27513<br>UNITED STATES |
| WESTERN INSTITUTIONAL REVIEW BOARD<br>1019 39th Ave S.E, Ste 120<br>Puyallup, WA 98374<br>UNITED STATES |
| Copernicus Group Institutional Review Board<br>5000 CentreGreen Way, Ste 200<br>Cary, NC 27513<br>UNITED STATES |
| Copernicus Group Institutional Review Board<br>5000 CentreGreen Way, Ste 200<br>Cary, NC 27513<br>UNITED STATES |
| Copernicus Group Institutional Review Board<br>5000 CentreGreen Way, Ste 200<br>Cary, NC 27513<br>UNITED STATES |
| Kaiser Permanente Northern California Institutional Review Board<br>1800 Harrison St, 10th Fl<br>Oakland, CA 94612<br>UNITED STATES |
| Copernicus Group Institutional Review Board<br>5000 CentreGreen Way, Ste 200<br>Cary, NC 27513<br>UNITED STATES |
| Copernicus Group IRB<br>5000 CentreGreen Way, Ste 200<br>Cary, NC 27513<br>UNITED STATES |
| Copernicus Group Institutional Review Board<br>5000 CentreGreen Way, Ste 200<br>Cary, NC 27513<br>UNITED STATES |
| Copernicus Group IRB<br>5000 Centregreen Way, Ste 200<br>Cary, NORTH CAROLINA 27513<br>UNITED STATES |
| Copernicus Group Institutional Review Board<br>5000 CentreGreen Way, Ste 200<br>Cary, NC 27513<br>UNITED STATES |
| Copernicus Group Institutional Review Board<br>5000 CentreGreen Way, Ste 200<br>Cary, NC 27513<br>UNITED STATES |
| Copernicus Group Institutional Review Board<br>5000 CentreGreen Way, Ste 200<br>Cary, NC 27513<br>UNITED STATES |
| Copernicus Group IRB<br>5000 Centregreen Way, Ste 200<br>Cary, NORTH CAROLINA 27513<br>UNITED STATES |
| Copernicus Group Institutional Review Board |

| |
|---|
| 5000 CentreGreen Way, Ste 200<br>Cary, NC 27513<br>UNITED STATES |
| WESTERN INSTITUTIONAL REVIEW BOARD<br>1019 39th Ave SE, Ste 120<br>Puyallup, WASHINGTON 98374-2115<br>UNITED STATES |
| WESTERN INSTITUTIONAL REVIEW BOARD<br>1019 39th Ave S.E, Ste 120<br>Puyallup, WA 98374<br>UNITED STATES |
| Western Institutional Review Board<br>1019 39th Ave SE, Ste 120<br>Puyallup, WASHINGTON 98374<br>UNITED STATES |
| Copernicus Group IRB<br>5000 CentreGreen Way, Ste 200<br>Cary, NORTH CAROLINA 27513<br>UNITED STATES |
| Copernicus Group Institutional Review Board<br>5000 CentreGreen Way, Ste 200<br>Cary, NC 27513<br>UNITED STATES |
| Copernicus Group Institutional Review Board<br>5000 CentreGreen Way, Ste 200<br>Cary, NC 27513<br>UNITED STATES |
| WCG IRB<br>1019 39th Ave Se, Ste 120<br>Puyallup, WASHINGTON 98374<br>UNITED STATES |
| Copernicus Group IRB<br>5000 Centregreen Way, Ste 200<br>Cary, NORTH CAROLINA 27513<br>UNITED STATES |
| Copernicus Group Institutional Review Board<br>5000 CentreGreen Way, Ste 200<br>Cary, NC 27513<br>UNITED STATES |
| Copernicus Group IRB<br>5000 Centregreen Way, Ste 200<br>Cary, NORTH CAROLINA 27513<br>UNITED STATES |
| Copernicus Group Institutional Review Board<br>5000 CentreGreen Way, Ste 200<br>Cary, NC 27513<br>UNITED STATES |
| Copernicus Group Institutional Review Board<br>5000 CentreGreen Way, Ste 200<br>Cary, NC 27513<br>UNITED STATES |
| Copernicus Group Institutional Review Board<br>5000 CentreGreen Way, Ste 200<br>Cary, NC 27513<br>UNITED STATES |

| |
|---|
| Copernicus Group Institutional Review Board<br>5000 CentreGreen Way, Ste 200<br>Cary, NC 27513<br>UNITED STATES |
| Copernicus Group IRB<br>5000 CentreGreen Way, Ste 200<br>Cary, NORTH CAROLINA 27513<br>UNITED STATES |
| Copernicus Group Institutional Review Board<br>5000 CentreGreen Way, Ste 200<br>Cary, NC 27513<br>UNITED STATES |
| Copernicus Group Institutional Review Board<br>5000 CentreGreen Way, Ste 200<br>Cary, NC 27513<br>UNITED STATES |
| Copernicus Group Institutional Review Board<br>5000 CentreGreen Way, Ste 200<br>Cary, NC 27513<br>UNITED STATES |
| Lehigh Valley Health Network/Institutional Review Board/Research Participant Office<br>1255 S Cedar Crest Blvd, Ste 3200<br>Allentown, PA 18103<br>UNITED STATES |
| Copernicus Group Institutional Review Board<br>5000 CentreGreen Way, Ste 200<br>Cary, NC 27513<br>UNITED STATES |
| Copernicus Group Institutional Review Board<br>5000 Centregreen Way, Ste 200<br>Cary, NORTH CAROLINA 27513<br>UNITED STATES |
| Copernicus Group Institutional Review Board<br>5000 CentreGreen Way, Ste 200<br>Cary, NC 27513<br>UNITED STATES |
| Copernicus Group Institutional Review Board<br>5000 CentreGreen Way, Ste 200<br>Cary, NC 27513<br>UNITED STATES |
| Copernicus Group Institutional Review Board<br>5000 CentreGreen Way, Ste 200<br>Cary, NC 27513<br>UNITED STATES |
| Copernicus Group IRB<br>5000 Centregreen Way, Ste 200<br>Cary, NORTH CAROLINA 27513<br>UNITED STATES |
| Western Institutional Review Board<br>1019 39th Ave SE, Ste 200<br>Puyallup, WA 98374-2115<br>UNITED STATES |
| Indian Health Service National IRB<br>5600 Fishers Ln, MS 09E10D<br>Rockville, MARYLAND 20857 |

| |
|---|
| UNITED STATES |
| Johns Hopkins Bloomberg School of Public Health<br>615 N. Wolfe St, Rm E1100<br>Baltimore, MARYLAND 21205<br>UNITED STATES |
| Navajo Nation Human Research Review Board<br>Window Rock Blvd, Administration Bldg #2, Division of Health, P.O. Box 1390<br>Window Rock, ARIZONA 86515<br>UNITED STATES |
| Johns Hopkins Bloomberg School of Public Health<br>615 N. Wolfe St, Rm E1100<br>Baltimore, MARYLAND 21205<br>UNITED STATES |
| Navajo Nation Human Research Review Board<br>Window Rock Blvd, Administration Bldg #2, Division of Health, P.O. Box 1390<br>Window Rock, ARIZONA 86515<br>UNITED STATES |
| Johns Hopkins Bloomberg School of Public Health<br>615 N. Wolfe St, Rm E1100<br>Baltimore, MARYLAND 21205<br>UNITED STATES |
| Johns Hopkins Bloomberg School of Public Health<br>615 N. Wolfe St, Rm E1100<br>Baltimore, MARYLAND 21205<br>UNITED STATES |
| Navajo Nation Human Research Review Board<br>Window Rock Blvd, Administration Bldg #2, Division of Health, P.O. Box 1390<br>Window Rock, ARIZONA 86515<br>UNITED STATES |
| Yale University Human Research Protection Program (Human Investigation Committee)<br>25 Science Park, 3rd Fl, 150 Munson St<br>New Haven, CT 06520<br>UNITED STATES |
| Copernicus Group IRB<br>5000 Centregreen Way, Ste 200<br>Cary, NORTH CAROLINA 27513<br>UNITED STATES |
| Copernicus Group Institutional Review Board<br>5000 CentreGreen Way, Ste 200<br>Cary, NC 27513<br>UNITED STATES |
| Western Institutional Review Board<br>1019 39th Ave. SE, Ste 120<br>Puyallup, WA 98374<br>UNITED STATES |
| Copernicus Group Institutional Review Board<br>5000 CentreGreen Way, Ste 200<br>Cary, NC 27513<br>UNITED STATES |
| Copernicus Group Institutional Review Board<br>5000 CentreGreen Way, Ste 200<br>Cary, NC 27513<br>UNITED STATES |
| Copernicus Group Institutional Review Board<br>5000 Centregreen Way, Ste 200 |

| |
|---|
| Cary, NORTH CAROLINA 27513<br>UNITED STATES |
| Copernicus Group Institutional Review Board<br>5000 CentreGreen Way, Ste 200<br>Cary, NC 27513<br>UNITED STATES |
| Copernicus Group Institutional Review Board<br>5000 CentreGreen Way, Ste 200<br>Cary, NC 27513<br>UNITED STATES |
| Western Institutional Review Board<br>1019 39th Ave SE, Ste 120<br>Puyallup, WA 98374<br>UNITED STATES |
| Copernicus Group Institutional Review Board<br>5000 CentreGreen Way, Ste 200<br>Cary, NC 27513<br>UNITED STATES |
| Western Institutional Review Board<br>1019 39th Ave SE, Ste 120<br>Puyallup, WA 98374<br>UNITED STATES |
| Copernicus Group Institutional Review Board<br>5000 CentreGreen Way, Ste 200<br>Cary, NC 27513<br>UNITED STATES |
| Copernicus Group Institutional Review Board<br>5000 CentreGreen Way, Ste 200<br>Cary, NC 27513<br>UNITED STATES |
| Kaiser Permanente Northern California Institutional Review Board<br>1800 Harrison St, 10th Fl<br>Oakland, CA 94612<br>UNITED STATES |
